# Point-of-care C-reactive protein and Xpert MTB/RIF Ultra for tuberculosis screening and diagnosis in unselected antiretroviral therapy initiators: a prospective diagnostic accuracy study

**DOI:** 10.1101/2023.05.30.23290716

**Authors:** Byron WP Reeve, Gcobisa Ndlangalavu, Hridesh Mishra, Zaida Palmer, Happy Tshivhula, Loren Rockman, Selisha Naidoo, Desiree L Mbu, Charissa C Naidoo, Brigitta Derendinger, Gerhard Walzl, Stephanus T. Malherbe, Paul D van Helden, Fred C Semitala, Christina Yoon, Rishi K Gupta, Mahdad Noursadeghi, Robin M Warren, Grant Theron

## Abstract

**Background:** Tuberculosis (TB), a major cause of death in people living with HIV (PLHIV), remains challenging to diagnose. Diagnostic accuracy data are lacking for promising triage tests, such as C-reactive protein (CRP), and confirmatory tests, such as sputum and urine Xpert MTB/RIF Ultra (Ultra), and urine LAM, without prior symptom selection.

**Methods:** 897 PLHIV initiating antiretroviral therapy were consecutively recruited in settings with high TB incidence, irrespective of symptoms. Participants were offered sputum induction, with a liquid culture reference standard. First, we evaluated point-of-care CRP testing on blood, compared to the World Health Organization (WHO)-recommended four-symptom screen (W4SS) for triage (n=800). Second, we evaluated Xpert MTB/RIF Ultra (Ultra) versus Xpert MTB/RIF (Xpert) for sputum-based confirmatory testing (n=787), with or without sputum induction. Third, we evaluated Ultra and Determine LF-LAM for urine-based confirmatory testing (n=732).

**Findings:** CRP and number of W4SS symptoms had areas under the receiver operator characteristic curve of 0.78 (95% confidence interval 0.73, 0.83) and 0.70 (0.64, 0.75), respectively. For triage, CRP (≥10 mg/l) has similar sensitivity to W4SS [77% (68, 85) vs. 77% (68, 85); p>0.999] but higher specificity [64% (61, 68) vs. 48% (45, 52); p<0.001]; reducing unnecessary confirmatory testing by 138 per 1000 people and the number-needed-to-test from 6.91 (6.25, 7.81) to 4.87 (4.41, 5.51). Using sputum, which required induction in 31% (24, 39) of people, Ultra had higher sensitivity than Xpert [71% (61, 80) vs. 56% (46, 66); p<0.001] but lower specificity [98% (96, 100) vs. 99% (98, 100); p<0.001]. The proportion of people with ≥1 positive confirmatory result detected by Ultra increased from 45% (26, 64) to 66% (46, 82) when induction was done. Programmatically-done haemoglobin, triage test combinations, and urine tests showed comparatively worse performance.

**Interpretation:** Among ART-initiators in a high burden setting, CRP is a more specific triage test than W4SS. Sputum induction improves yield. Sputum Ultra is a more accurate confirmatory test than Xpert.

**Funding:** SAMRC (MRC-RFA-IFSP-01-2013), EDCTP2 (SF1401, OPTIMAL DIAGNOSIS), NIH/NIAD (U01AI152087).

**Research in context:** *Evidence before this study:* Novel triage and confirmatory tests are urgently needed for TB, especially in key risk groups like PLHIV. Many TB cases do not meet World Health Organization (WHO)-recommended four-symptom screen (W4SS) criteria despite accounting for significant transmission and morbidity. W4SS also lacks specificity, which makes onward referral of triage-positive people for expensive confirmatory testing inefficient and hampers diagnostic scale-up. Alternative triage approaches like CRP have promise, but have comparatively little data in ART-initiators, especially when done without syndromic preselection and using point-of-care (POC) tools. After triage, confirmatory testing can be challenging due to sputum scarcity and paucibacillary early-stage disease. Next generation WHO-endorsed rapid molecular tests (including Xpert MTB/RIF Ultra; Ultra) are a standard-of-care for confirmatory testing. However, there are no supporting data in ART-initiators, among whom Ultra may offer large sensitivity gains over predecessors like Xpert MTB/RIF (Xpert). The added value of sputum induction to augment diagnostic sampling for confirmatory testing is also unclear. Lastly, the performance of urine tests (Ultra, Determine LF-LAM) in this population requires more data.

*Added value of this study:* We evaluated repurposed and new tests for triage and confirmatory testing using a rigorous microbiological reference standard in a highly vulnerable high-priority patient population (ART-initiators) regardless of symptoms and ability to naturally expectorate sputum. We showed POC CRP triage is feasible, performs better than W4SS, and that combinations of different triage approaches offer no advantages over CRP alone. Sputum Ultra has superior sensitivity to Xpert; often detecting W4SS-negative TB. Furthermore, without induction, confirmatory sputum-based testing would not be possible in a third of people. Urine tests had poor performance. This study contributed unpublished data to systematic reviews and meta-analyses used by the WHO to inform global policy supporting use of CRP triage and Ultra in PLHIV.

*Implication of all the available evidence:* POC CRP triage testing is feasible and superior to W4SS and, together with sputum induction in people who triage CRP-positive should, after appropriate cost and implementation research, be considered for roll-out in ART-initiators in high burden settings. Such people should be offered Ultra, which outperforms Xpert.

## Introduction

Tuberculosis (TB) is the single biggest cause of death among people living with HIV (PLHIV). Such individuals are most immunocompromised prior to antiretroviral therapy (ART) initiation and, at HIV diagnosis, are already captured within HIV care cascades. ART initiation clinics represent a valuable opportunity to rapidly screen and test for TB. However, the point-of-care (POC) feasibility, accuracy, and potential utility of new and repurposed tools is unclear, especially in the absence of symptomatic pre-selection^1^.

The World Health Organization (WHO)-recommended four-symptom screen (W4SS) is used to identify people requiring confirmatory TB testing. However, in addition to being subjective, stigmatizing and poorly implemented^2, 3^, prevalence surveys show that most bacteriologically positive TB is in W4SS-negative individuals^4^. Consequently, the WHO emphasises that, as an alternative or adjunct to symptoms, an inexpensive and rapid triage test with ≥90% sensitivity and ≥70% specificity on accessible non-sputum specimens is needed to better focus relatively expensive confirmatory testing^5^.

C-reactive protein (CRP) is a blood biomarker with evidence to assist in TB diagnosis in PLHIV^6–11^. CRP platforms are commercially-available^12^, however, few are evaluated at POC^13^ or alongside other biomarkers like haemoglobin^14^. For the implementation of CRP triage, POC testing is required to enable real-time decision making within the same encounter that symptoms are ascertained. A systematic review and individual participant data meta-analysis, which focussed on aggregate data in PLHIV but not specifically ART-initiators, W4SS- negatives, or POC feasibility, showed CRP to have similar sensitivity vs. W4SS but higher specificity^15^. This meta-analysis informed a WHO guideline update, where CRP was recommended as an adjunct to W4SS triage for HIV-associated TB^16^.

CRP performance data in concert with the new Xpert MTB/RIF Ultra (Ultra) sputum test are also limited. Ultra has improved limit of detection for *Mycobacterium tuberculosis* complex (*Mtb*) versus its predecessor Xpert MTB/RIF (Xpert). Critically, this sensitivity increment is likely largest in people with paucibacillary early-stage disease^17^. Such individuals may not self-present to care due to TB symptoms (if any) but still do so, for example, to initiate ART. Ultra at this point could therefore represent a hitherto unavailable opportunity to detect TB early in a key risk group, however, there are limited data.

Importantly, individuals with early-stage TB disease and/or HIV may not be able to naturally expectorate sputum^18^. Sputum-scarce individuals are often excluded from rapid confirmatory testing – both programmatically and in research studies – because sputum induction is frequently unavailable in high burden primary care settings. Diagnostic reference standards are also challenging in people from whom a sputum was not retrieved and sputum-scarce people hence represent a source of missed cases not included in most sensitivity calculations. Furthermore, the proportion of people who would have needed induction is seldom reported, meaning that studies that include only sputum expectorators overestimate real-world test performance. Lastly, due to these and other challenges associated with sputum^19^, it is important to evaluate confirmatory tests on easily accessible fluids like urine.

We therefore sought to evaluate the performance of novel triage and confirmatory testing approaches among PLHIV who were unselected based on symptoms and visiting a facility in Cape Town, South Africa to start ART. For triage, we compared POC CRP to W4SS as the current standard-of-care. For confirmatory testing, we evaluated sputum tests (Xpert, Ultra), the incremental diagnostic yield of sputum induction, and urine tests (Ultra, LF-LAM).

## Methods

### Study population and flow

897 adults (≥18 years) newly diagnosed with HIV and referred to Kraaifontein Community Health Centre to start ART were prospectively enrolled (15 March 2017 - 17 March 2020) without symptom criteria and evaluated at a single visit (**Figure 1**). Participants were excluded if they had received any TB treatment within the last two months, were of unknown treatment status, or did not consent.

**Figure 1.**
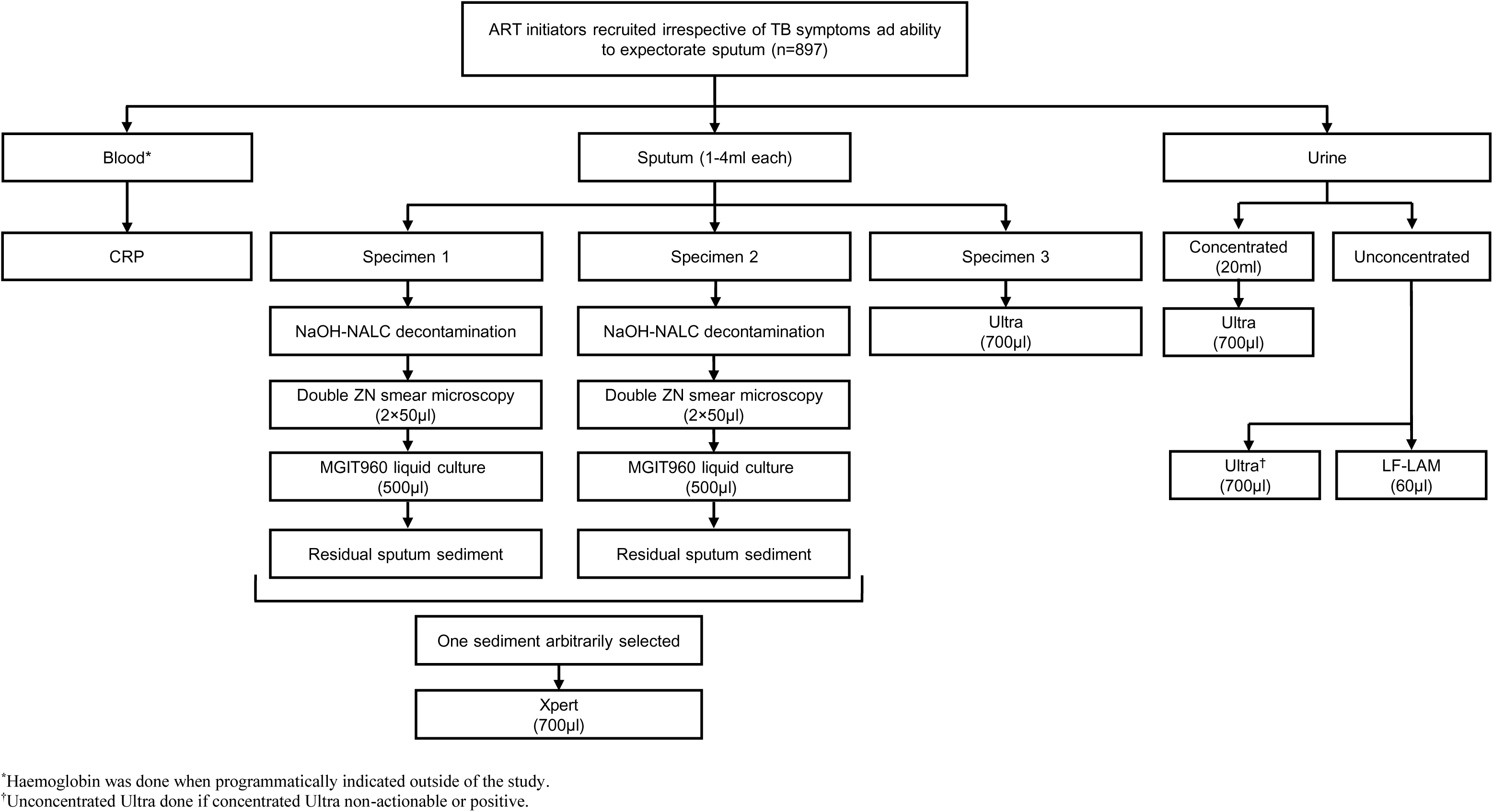
Study profile showing participant enrolment, specimen collection and processing, and tests done. Abbreviations: ART, antiretroviral treatment; CRP, C-reactive protein; LF-LAM, Determine TB LAM Ag test (LF-LAM); MGIT960, Mycobacterial Growth Indicator Tube; TB, tuberculosis; Ultra, Xpert MTB/RIF Ultra; Xpert, Xpert MTB/RIF; ZN, Ziehl– Neelsen.

### Ethics

This study was approved by the Stellenbosch University Faculty of Health Sciences Research Ethics Committee (N14/10/136) and the Western Cape Department of Health, South Africa (WC_2016RP38_944), and is on Clinicaltrials.gov (NCT03187964).

### W4SS, CRP, and haemoglobin

People required at least one W4SS symptom (current cough, fever, night sweats or weight loss) to be positive^16^. Four-to-five drops of capillary blood were used by a health worker for POC CRP measurement (iChromaII platform, Boditech, South Korea) (**Supplement)**. CD4 counts were measured outside the study but captured (counts more than three months before or after recruitment were excluded) as well as haemoglobin (when medically indicated by routine staff per local guidelines^20^). CRP positivity thresholds of ≥5 or ≥10 mg/l (CRP_5_, CRP_10_) and <10 g/dl haemoglobin were pre-selected based on literature^10, 15^. Different combinations of triage tests (“algorithms” defined in **Table 3**) were evaluated.

### Specimen collection and bacteriological tests

Three sputa (≥1 ml each) were required from participants, who first attempted expectoration before induction was offered (whether specific individual sputa made were expectorated or induced was, due to a database error, only successfully recorded for the last 158 participants).

Induction used a nebuliser (Ultrasonic Hospital Grade WH-802, Hitech Therapy, South Africa) with 5% NaCl (Ysterplaat Medical Supplies, South Africa) for 7-10 min^18^. Two sputa were arbitrarily selected to each undergo 1% NaOH-NALC decontamination, Ziehl-Neelsen (ZN) smear microscopy, and Mycobacteria Growth Indicator Tube (MGIT) 960 culture. The other sputum underwent Ultra testing and Xpert was done on the sputum sediment remnant remaining after culture inoculation. The **Supplement** has further information on specimen storage, sputum, urine, and isolate testing. Study staff had access to all clinical, index test, and reference standard data. Sputum Ultra and microscopy (both available ≤48 h), culture and MTBDR*plus* results (typically available within 35 days) were reported for potential participant management to the programme. No adverse events occurred.

### Reference standard

If at least one culture was *Mtb-*positive, participants were classified as having TB. Participants with negative culture(s) did not have TB.

### Statistical analyses

Data were captured on RedCap^21^. Head-to-head analyses (only people with actionable results from the relevant tests) are presented unless stated otherwise. Sensitivity, specificity, and predictive values for tests and algorithms were calculated using two-by-two tables with 95% confidence intervals (binomial proportion method). Yield was calculated as, of the people who had an index test attempted, those with a positive result [sputum Ultra, sputum Xpert, urine Ultra (concentrated or unconcentrated), urine LF-LAM]. People who did not have a test attempted, due to human error for example, were excluded from yield calculations and non-actionable results (not positive or negative^22, 23^; **Supplement**) were included. Statistical tests included McNemar’s chi-squared, Mann-Whitney, two-sample proportion, Kruskal-Wallis and Spearman’s coefficient using Stata (version 15; StataCorp) and GraphPad Prism (version 7.0; GraphPad Software). Culture-negative people who were urine Ultra-positive had medical record information up to one-year post-recruitment requested to evaluate if they were later sputum confirmatory test-positive. We followed STARD analysis and reporting criteria^24^.

## Results

### Patient characteristics

12% (107/897) of the total population was culture-positive [34% (36/107) positive by one culture]. This comprised 13% (104/800) of people in our comparison of CRP and W4SS triage tests, 13% (104/787) in our comparison of sputum confirmatory tests, and 13% (97/732) in our urine test comparison (**Figure 2A-C)**. Patient characteristics, including W4SS, TB, and smear statuses, are in **Table 1**. 25% (26/104) of people with TB were W4SS-negative.

**Figure 2.**
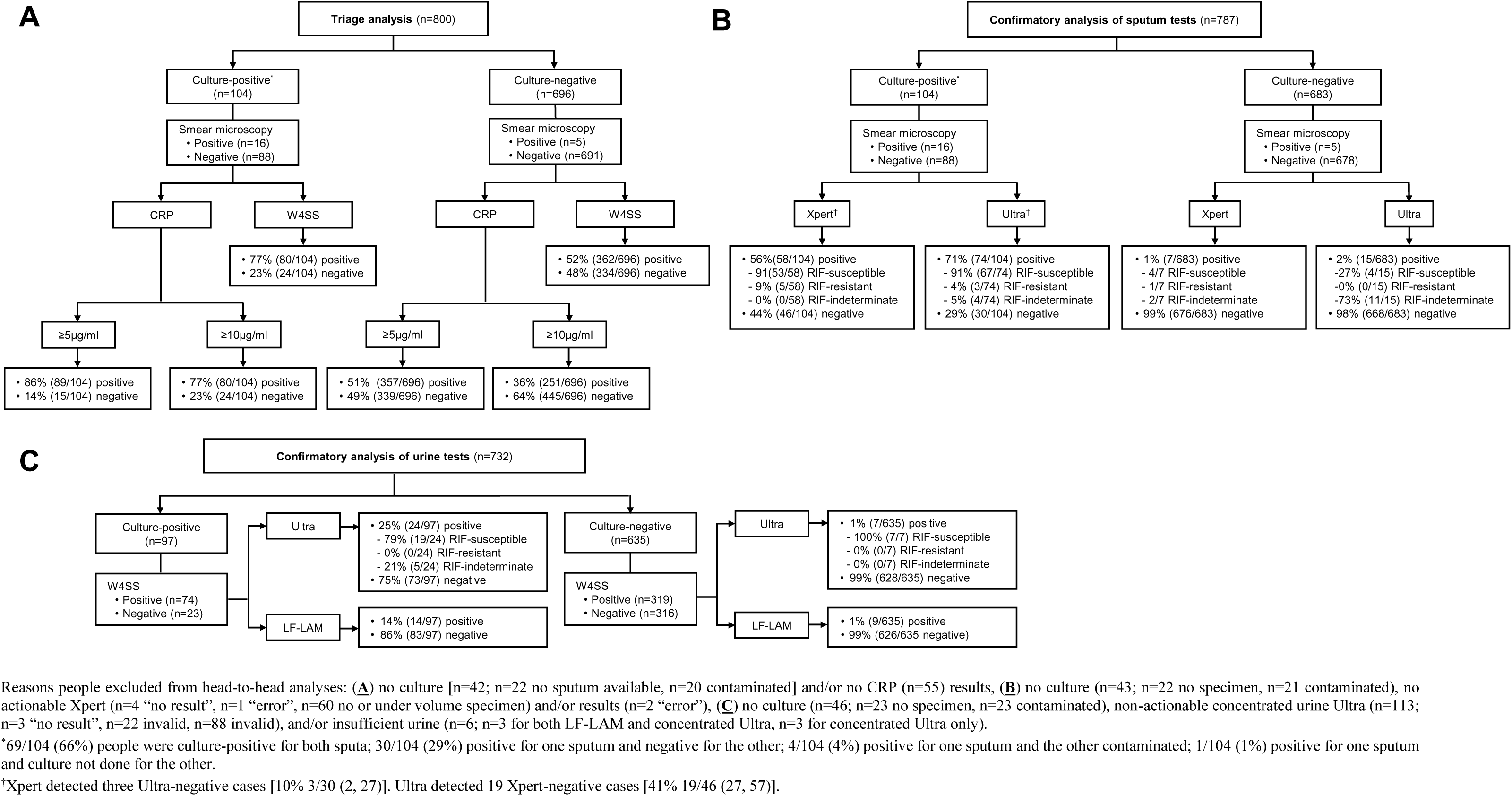
Flow diagrams showing the number of people in head-to-head comparisons of (**A**) triage (CRP, W4SS), (**B**) confirmatory (Xpert, Ultra), and (**C**) urine (concentrated Ultra, LF-LAM) tests. CRP correctly classified more people without TB compared to W4SS, Ultra detected more TB than Xpert on sputum and, on concentrated urine, more TB than LF-LAM. Abbreviations: CRP, C-reactive protein; LF-LAM, Determine TB LAM Ag ; POC, point of care; RIF, rifampicin; Ultra, Xpert MTB/RIF Ultra; W4SS, WHO-recommended four-symptom screen; Xpert; Xpert MTB/RIF.

**Table 1.** Demographic and clinical characteristics by W4SS, culture, and smear statuses. Compared to W4SS-positives, W4SS-negatives were more likely to be younger, female, non-smokers, culture-negative, have lower CRP but higher CD4 and haemoglobin levels. W4SS-positives were also more likely to have TB, be able to naturally expectorate sputum and, in those with TB, greater sputum mycobacterial load. Culture-positives were, compared to culture-negatives, more likely to be older, of mixed ancestry, smokers, have lower CD4 counts and haemoglobin, and higher CRP levels. Data are median (IQR) or n (%).

|  | Overall<br>n=787 | W4SS status |  | Culture and smear status |  |  |  |
| --- | --- | --- | --- | --- | --- | --- | --- |
|  |  | Positive<br>430/787<br>(55) | Negative<br>357/787<br>(45) | Culture-positive |  |  | Culture-negative<br>683/787<br>(87) |
|  |  |  |  | All<br>104/787<br>(13) | Smear-positive<br>16/104<br>(15) | Smear-negative<br>88/104<br>(85) |  |
| Demographic |  |  |  |  |  |  |  |
| Age, years | 32<br>(26-39) | 33<br>(28-40) | 31<br>(25-37)<br>p<0.001* | 34<br>(29-39) | 37<br>(33-42) | 34<br>(29-39)<br>p=0.170<sup>‡</sup> | 31<br>(26-38)<br>p=0.010** |
| Female | 463/787<br>(59) | 233/430<br>(54) | 230/357<br>(64)<br>p<0.001* | 43/104<br>(41) | 4/16<br>(25) | 39/88<br>(44)<br>p=0.149<sup>‡</sup> | 420/683<br>(61)<br>p<0.001** |
| Ethnicity<sup>‡</sup> |  |  |  |  |  |  |  |
| Mixed ancestry | 158/787<br>(21) | 93/430<br>(22) | 70/357<br>(20)<br>p=0.486* | 31/104<br>(30) | 4/16<br>(25) | 27/88<br>(31)<br>p=0.648<sup>‡</sup> | 132/683<br>(19)<br>p=0.014** |
| Black | 618/787<br>(79) | 334/430<br>(78) | 284/357<br>(80)<br>p=0.009* | 72/104<br>(69) | 11/16<br>(69) | 61/88<br>(69)<br>p=0.964<sup>‡</sup> | 546/683<br>(80)<br>p=0.013** |
| Tobacco smoker (past or current) | 372/787<br>(47) | 239/430<br>(56) | 133/357<br>(37)<br>p<0.001* | 64/104<br>(62) | 9/16<br>(56) | 55/88<br>(63)<br>p=0.636<sup>‡</sup> | 308/683<br>(45)<br>p<0.001** |
| Clinical |  |  |  |  |  |  |  |
| Number of W4SS symptoms (0-4) | 1<br>(0-2) | 2<br>(1-2) | - | 2<br>(1-3) | 3<br>(2-3) | 2<br>(1-3)<br>p=0.171<sup>‡</sup> | 1<br>(0-2)<br>p<0.001** |
| Able to expectorate at least one ≥1 ml sputum | 80/121<sup>†</sup><br>(66) | 47/79<br>(59) | 33/42<br>(79)<br>p=0.035* | 16/24<br>(67) | 3/5<br>(60) | 13/19<br>(68)<br>p=0.722<sup>‡</sup> | 64/97<br>(66)<br>p=0.949** |
| Culture-positive | 104/787<br>(13) | 78/430<br>(18) | 26/357<br>(7)<br>p<0.001* | 104/104<br>(100) | 16/16<br>(100) | 88/88<br>(100)<br>p>0.999<sup>‡</sup> | - |
| TTP (days) | 13<br>(9-18) | 12<br>(9-15) | 15<br>(12-21)<br>p=0.013* | 13<br>(9-18) | 6<br>(5-7) | 13<br>(11-19)<br>p<0.001<sup>‡</sup> | - |
| Previous TB | 112/787<br>(14) | 69/430<br>(16) | 43/357<br>(12)<br>p=0.110* | 16/104<br>(15) | 3/16<br>(19) | 13/88<br>(15)<br>p=0.685<sup>‡</sup> | 96/683<br>(14)<br>p=0.718** |
| CD4 (cells/μl) | 299<br>(170-483) | 263<br>(110-428) | 348<br>(227-521)<br>p<0.001* | 193<br>(60-285) | 199<br>(45-257) | 192<br>(61-285)<br>p=0.896<sup>‡</sup> | 326<br>(197-506)<br>p<0.001** |
| CRP (mg/l) | 7<br>(3-34) | 14<br>(3-77) | 4<br>(3-11)<br>p<0.001* | 72<br>(14-150) | 125<br>(101-286) | 45<br>(10-139)<br>p<0.001<sup>‡</sup> | 6<br>(3-20)<br>p<0.001** |
| Haemoglobin (g/dl) | 13<br>(12-14) | 13<br>(11-14) | 14<br>(12-15)<br>p<0.001* | 12<br>(10-13) | 10<br>(10-13) | 12<br>(10-13)<br>p=0.311<sup>‡</sup> | 13<br>(12-14)<br>p<0.001** |

<sup>a</sup>N are shown for people included in the Ultra and Xpert head-to-head comparison. Within row p-values are for comparisons by <sup>\*</sup>W4SS, <sup>‡</sup>smear, or <sup>\*\*</sup>culture statuses. <sup>‡</sup>Eight people self-reported as “other ethnicity”. <sup>†</sup>121 people had whether sputum was expectorated or induced recorded. <sup>§</sup>Negative values are days before recruitment, positives days after. Missing or not done: CRP (n=171; n=54 not done; n=117 outside detection range, see **Supplement**), CD4 (n=4 not done; n=47 older than three months), haemoglobin (n=50 not captured). Abbreviations: CRP, C-reactive protein; IQR, interquartile range; TB, tuberculosis; TTP, time-to-positivity; W4SS, WHO-recommended four-symptom screen.

### Triage tests

#### Individual biomarker sensitivity and specificity

Cough (any), ≥2-week cough, and W4SS had a sensitivity and specificity of 53% [95% confidence interval (CI) 43, 63] and 73% (70, 76), 42% (33, 53) and 83% (81, 86), and 77% (68, 85) and 48 (45, 52), respectively **(Table 2)**. POC CRP testing was feasible: 1% (5/835) of tests were non-actionable, all resolved upon re-testing, and all prospective results were generated within three minutes of finger prick. Sensitivity and specificity for CRP_5_ were 86% (78, 92) and 49% (45, 53), and for CRP_10_ were 77% (68, 85) and 64% (61, 68). Sensitivity at both thresholds was decreased in people with CD4 counts >350 vs. ≤350 cells/µl and specificity increased. When comparisons were limited to smear-negatives or included people that did not have head-to-head data, similar trends were observed (**Supplementary Tables 1-4**). Haemoglobin had 31% (22, 43) sensitivity and 88% (86, 91) specificity, displaying similar trends to CRP across CD4 count strata (**Supplementary Table 5**).

**Table 2.**
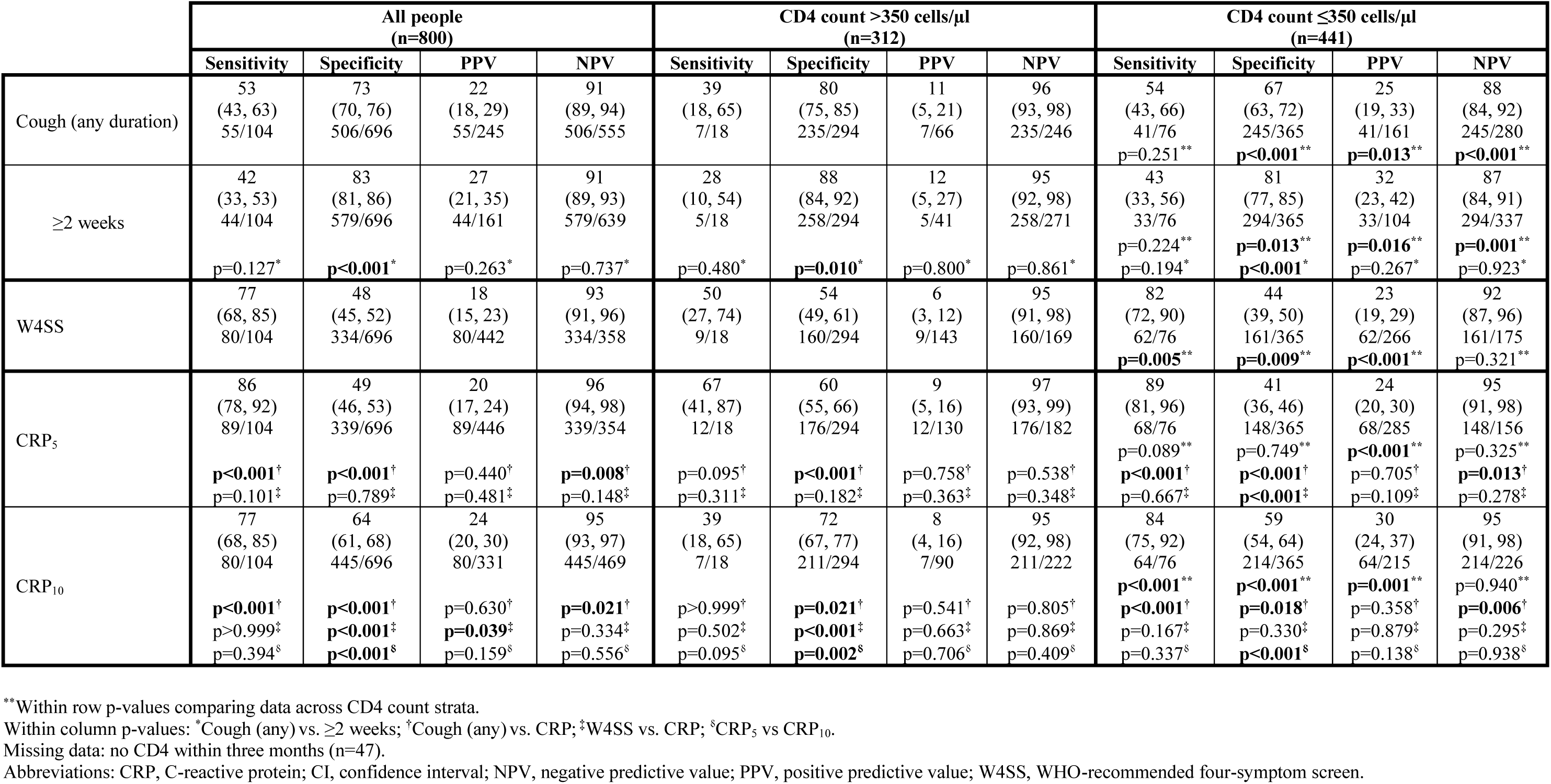
Head-to-head diagnostic accuracy of cough, W4SS, and CRP stratified by CD4 count. Cough (any or ≥2 weeks) had decreased sensitivity vs. W4SS but W4SS had sub-optimal specificity. In people with CD4 count >350 cells/µl, only the ≥ 10 mg/l CRP (CRP_10_) threshold (not ≥ 5 mg/l; CRP_5_) had increased specificity vs. W4SS whereas in people CD4 counts ≤350 cells/µl CRP_5_ and not CRP_10_ had increased specificity vs. W4SS. CRP_10_ had increased specificity CRP_5_ within each CD4 count stratum without a sensitivity decrement. Data are %, 95% CI, and n/N.

#### Individual biomarker receiver operating characteristic curves

W4SS and CRP receiver operating characteristic (ROC) curves are in **Figure 3** (haemoglobin in **Supplementary Figure 1**). Areas under the ROC curves (AUROCs) were 0.78 (0.73, 0.83) for CRP, 0.70 (0.64, 0.75) for W4SS, and 0.70 (0.64, 0.75) for haemoglobin. Among people with CD4 counts ≤350 cells/µl, higher AUROCs occurred for each biomarker vs. those with >350 cells/µl. AUROCs were also higher for W4SS-positives vs. -negatives. We next assessed biomarker performance under different scenarios and identified corresponding thresholds. At a rule-out threshold with sensitivity prioritised (∼95%), CRP (>3 mg/l; CRP_3_) sensitivity was similar vs. the WHO-recommended CRP_5_ [89% (83, 94) vs. 86% (78, 92)] and specificity diminished [39% (36, 42) vs. 49% (45, 53)], suggesting CRP_3_ offers small sensitivity improvements at large specificity costs. CRP and haemoglobin had small variations in rule-out thresholds across CD4 cell count and W4SS strata. Sensitivities and specificities under rule-in and Youden’s index scenarios are in **Supplementary Table 6**.

**Figure 3.**
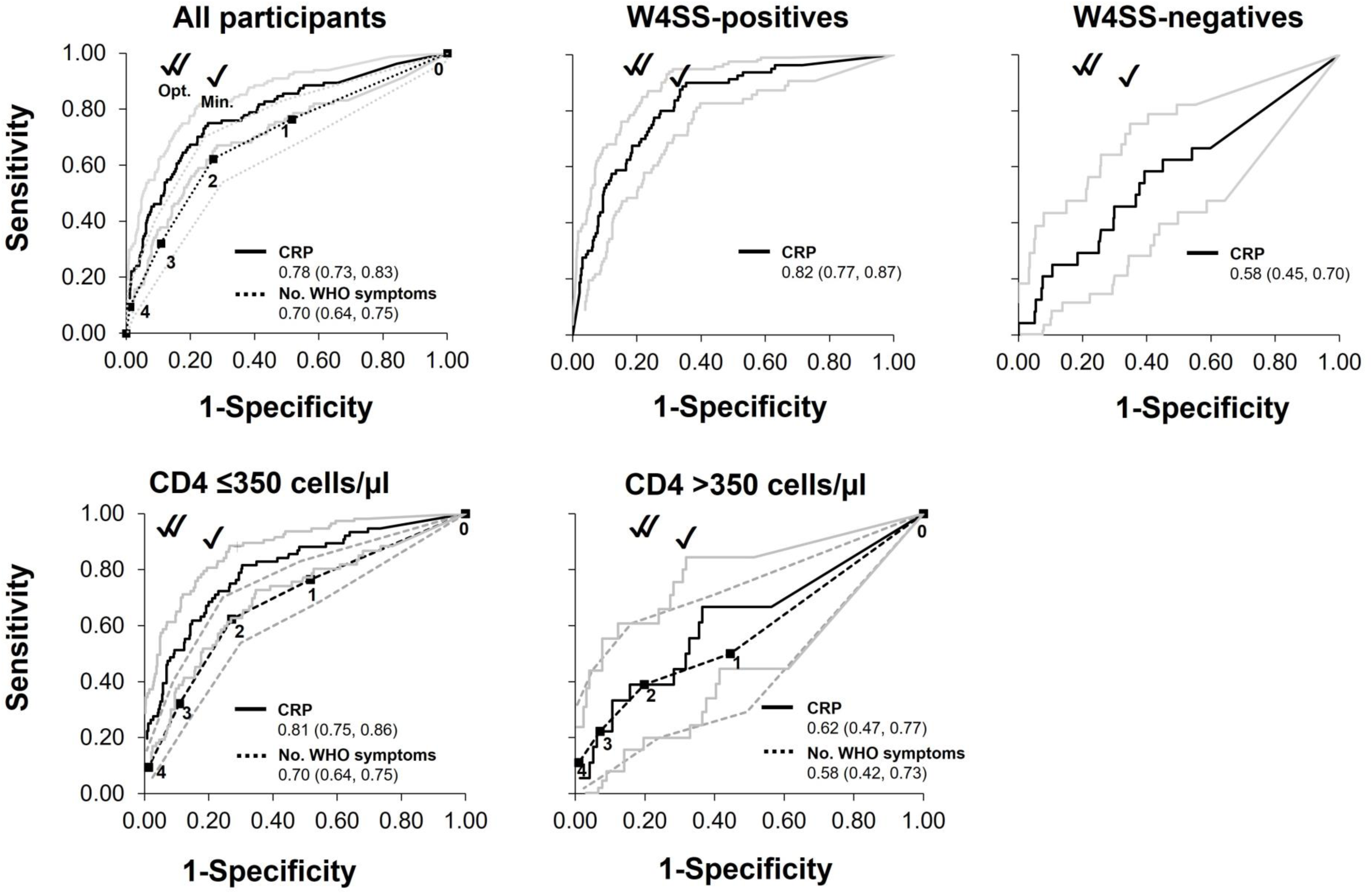
ROCs of CRP and number of W4SS in (**A**) all ART initiators, (**B, C**) stratified by W4SS-status, and (**D, E**) by CD4 cell counts. CRP had higher AUCROC in W4SS-positives than -negatives and in patients with lower than higher CD4 cell counts (as did W4SS). CRP was the only test for which CI overlapped with the WHO TPP minimal sensitivity and specificity target, and this was only in people with advanced disease (W4SS-positive or low CD4 count). AUROCs with 95% CIs with double and single ticks approximating the optimum and minimum WHO TPP sensitivity and specificity, respectively are shown. The number of symptoms are shown. Abbreviations: CRP, C-reactive protein; Opt., optimal WHO TPP; Min., minimum; WHO TPP; WHO TPP, World Health Organization target product profile; W4SS, WHO-recommended four-symptom screen.

#### Test combinations

Each biomarker’s sensitivity decreased in W4SS-negatives vs. -positives whereas specificity increased **(Supplementary Tables 1, 3)**. Triage test combinations were assessed as part of “algorithms” in **Table 3**. Algorithm 3 (triage positive if W4SS-positive and CRP_10_-positive) had similar sensitivity [71% (60, 81)] to W4SS [77% (68, 85)] and CRP_10_ [77% (68, 85)] individually, but specificity improved vs. each biomarker [77% (74, 81) vs. 48% (45, 52) for W4SS or vs 64% (61, 68) for CRP_10_] (**Table 2, 3**). Other algorithms, including CRP in W4SS- negatives, had similar sensitivities and generally diminished specificities versus their individual components **(Figure 4A)**. We did not include haemoglobin as it was in a programmatically-selected subset.

**Table 3.**
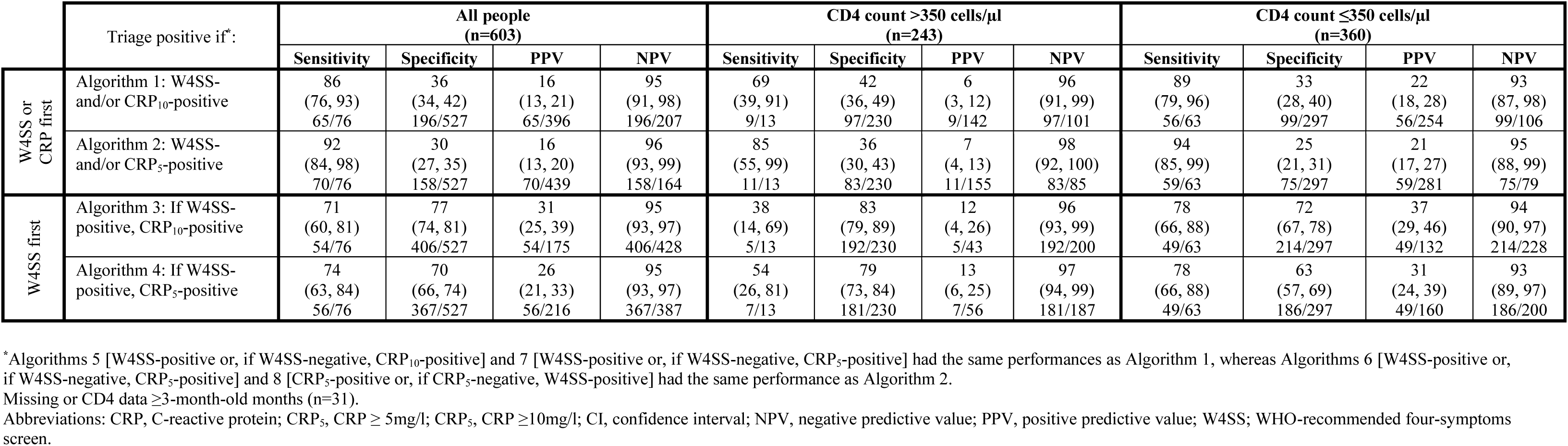
Diagnostic accuracy of parallel and sequential algorithms involving W4SS and CRP (stratified by CD4 cell count). A sequential W4SS-CRP approach (Algorithm 3) resulted in a more balanced sensitivity and specificity whereas a parallel approach (Algorithm 1) resulted in higher sensitivity but lower specificity. Each algorithm’s sensitivity increased at lower CD4 counts compared to higher counts while specificity decreased. People who, in the second column, do not meet the definition of triage-positive are triage-negative. Data are %, 95% CI, and n/N.

**Figure 4.**
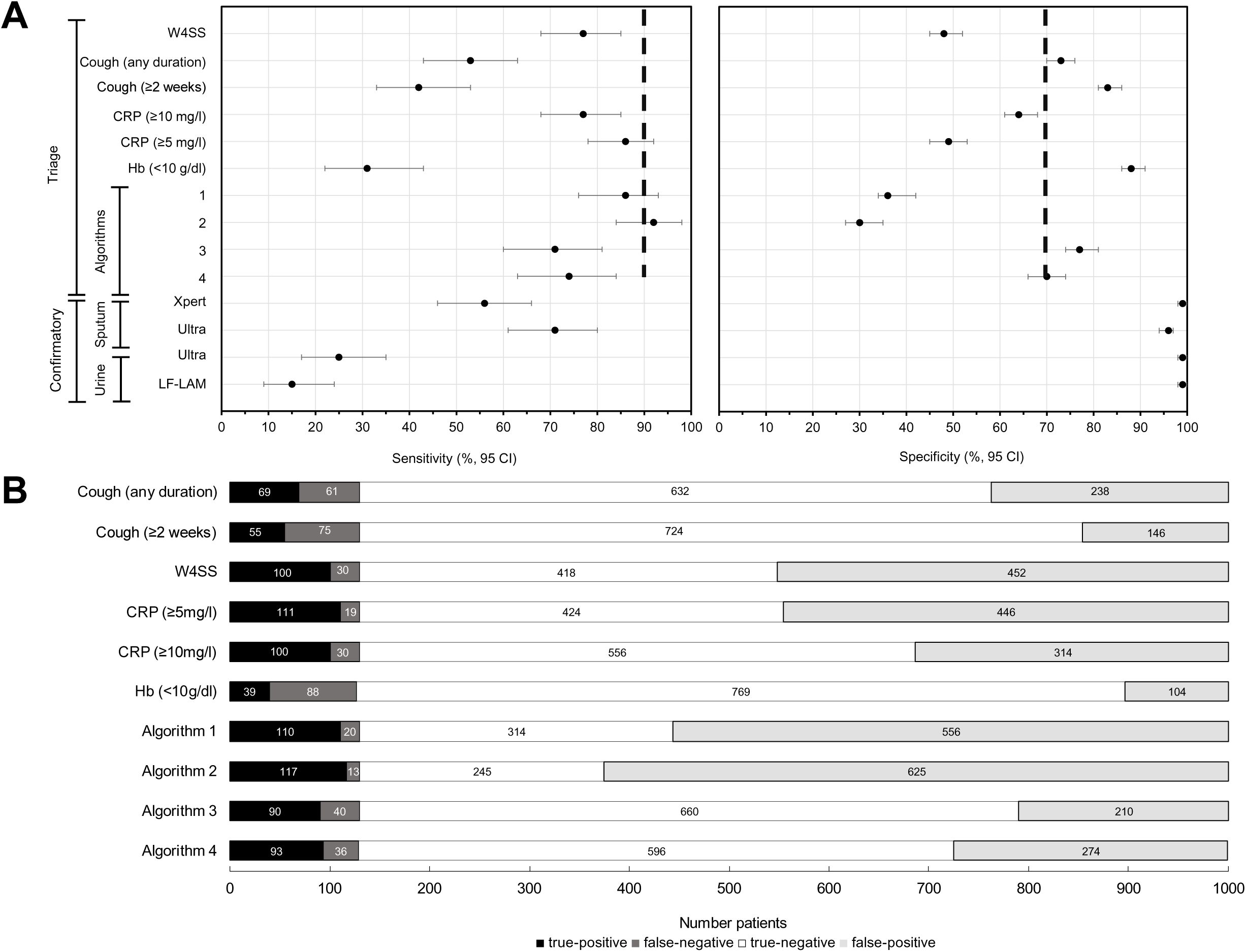
Summaries of triage and confirmatory test performance. **(A)** Forrest plots comparing sensitivity and specificity point estimates (with 95% CIs) of triage tests and algorithms as well as sputum and urine confirmatory tests. Black dashed vertical lines indicate WHO TPP estimates. **(B)** Effect of different triage tests and algorithms (defined in **Table 3**) on participant classification, showing CRP_10_ alone or in combination with W4SS (Algorithms 3 and 4) to result in fewer onward unnecessary referrals (false-positives). Abbreviations: CRP, C-reactive protein; Hb, haemoglobin; LF-LAM, Determine TB LAM Ag test; TB, tuberculosis; Ultra, Xpert MTB/RIF Ultra; W4SS, WHO-recommended four-symptom screen; WHO, World Health Organization; WHO TTP, WHO target product profile; Xpert, Xpert MTB/RIF.

#### Comparisons to WHO TPP sensitivity and specificity criteria

No point estimate achieved the minimum WHO triage test sensitivity target (90%), however, the CRP_5_ and Algorithm 2 95% CIs [86% (78, 92) and 92 (84, 98), respectively] overlapped 90%. The Algorithms 3 and 4 specificity estimate [77% (74, 81) and 70 (66, 74), respectively)] overlapped with the WHO minimum specificity estimate (70%).

#### Effect of triage tests and algorithms on confirmatory tests required

The effect of individual tests and algorithms on the number needing downstream confirmatory testing (number needed to test, NNT) at different prevalences is in **Figure 5**. Haemoglobin resulted in the smallest number of unnecessary referrals or false-positives, but this was at the expense of few people with TB correctly referred (true-positives). CRP_10_ reduced unnecessary testing more than W4SS, and this was enhanced further by a combination strategy also involving W4SS (Algorithms 3 and 4) but this resulted in fewer people with TB correctly referred. Briefly, CRP_10_ use would, versus W4SS, reduce Ultra NNT from 6.91 (6.25, 7.81) to 4.87 (4.41, 5.51). Improvements offered by CRP10 vs. W4SS remained relatively consistent across prevalences, with the NNT of triage strategies plateauing beyond 30% prevalence.

**Figure 5.**
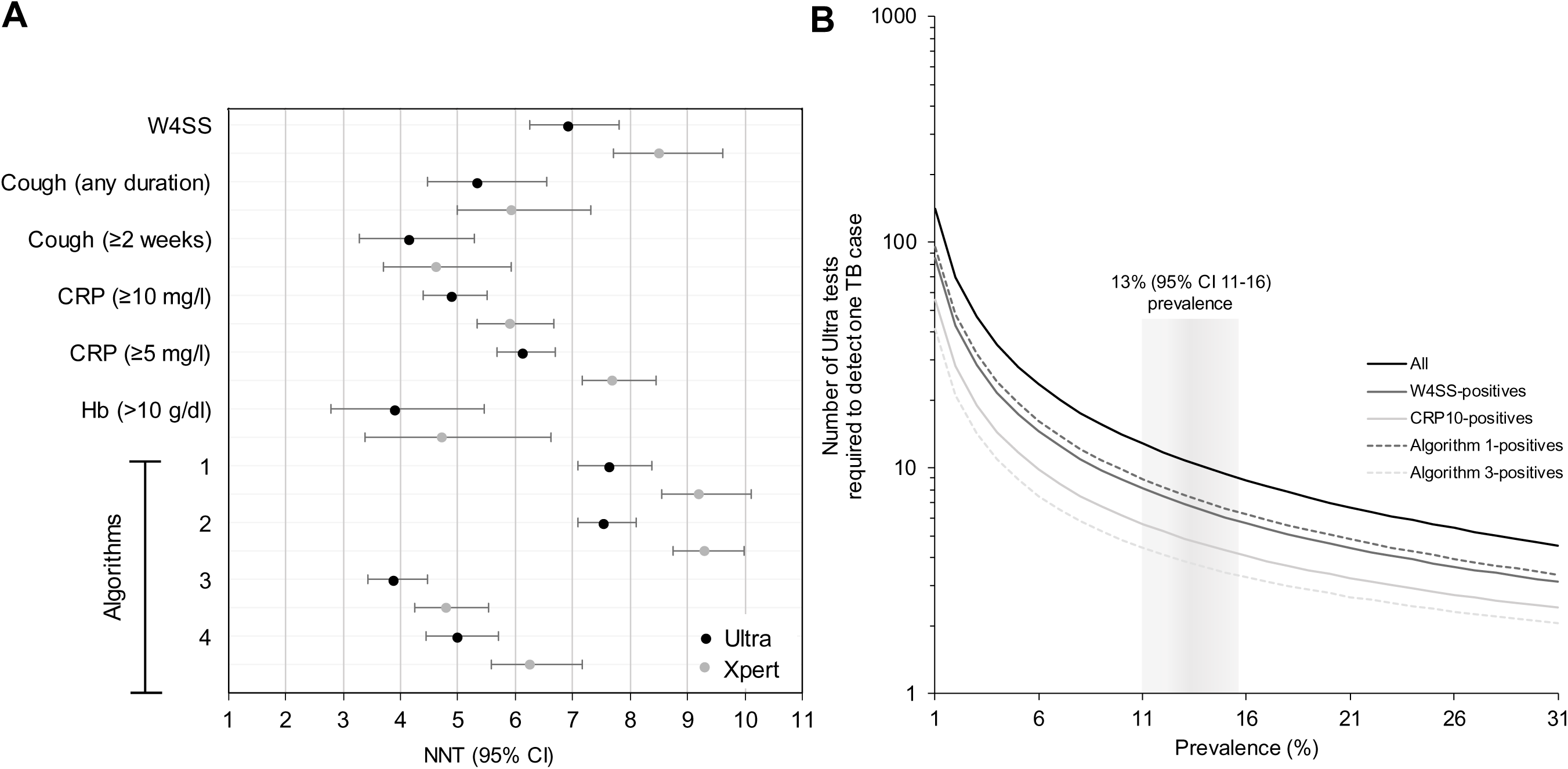
NNT for Ultra and Xpert to detect one culture-positive TB case when different triage methods are used in **(A)** our cohort and **(B)**, for Ultra only, modelled against different TB prevalences vs. an Ultra-in-all scenario. Use of CRP_10_ would result in a TB case correctly detected every five rather than, as for W4SS, every seven people. Other triage methods had lower NNTs but these would be offset by diminished sensitivities. In **B**, not all triage methods are shown and the grey column shows our prevalence. Abbreviations: CRP, C-reactive protein; Hb, haemoglobin; NNT, number needed to test; Ultra, Xpert MTB/RIF Ultra; W4SS, World Health Organization-recommended four symptom screen; Xpert, Xpert MTB/RIF.

### Sputum Ultra and Xpert diagnostic accuracy

#### Overall

Ultra and Xpert non-actionable results rates were low [1% (5/811), 1% (2/883)]. Ultra had, vs. Xpert, higher sensitivity [71% (61, 80) vs. 56% (46, 66) overall, 67% (56, 77) vs. 50% (39, 61) in smear-negatives], and lower specificity [96% (94, 97) vs. 99% (98, 100)] (**Table 4**; **Supplementary Table 7**). The NNT for Ultra to detect a culture-positive person was less than Xpert [4.87 (4.41, 5.51) vs. 5.90 (5.34. 6.67) in CRP_10_-positives, for example] Forest plots that include all confirmatory tests are in **Figure 4A** and Euler diagrams showing overlap between confirmatory tests are in **Supplementary Figure 2.**

**Table 4.**
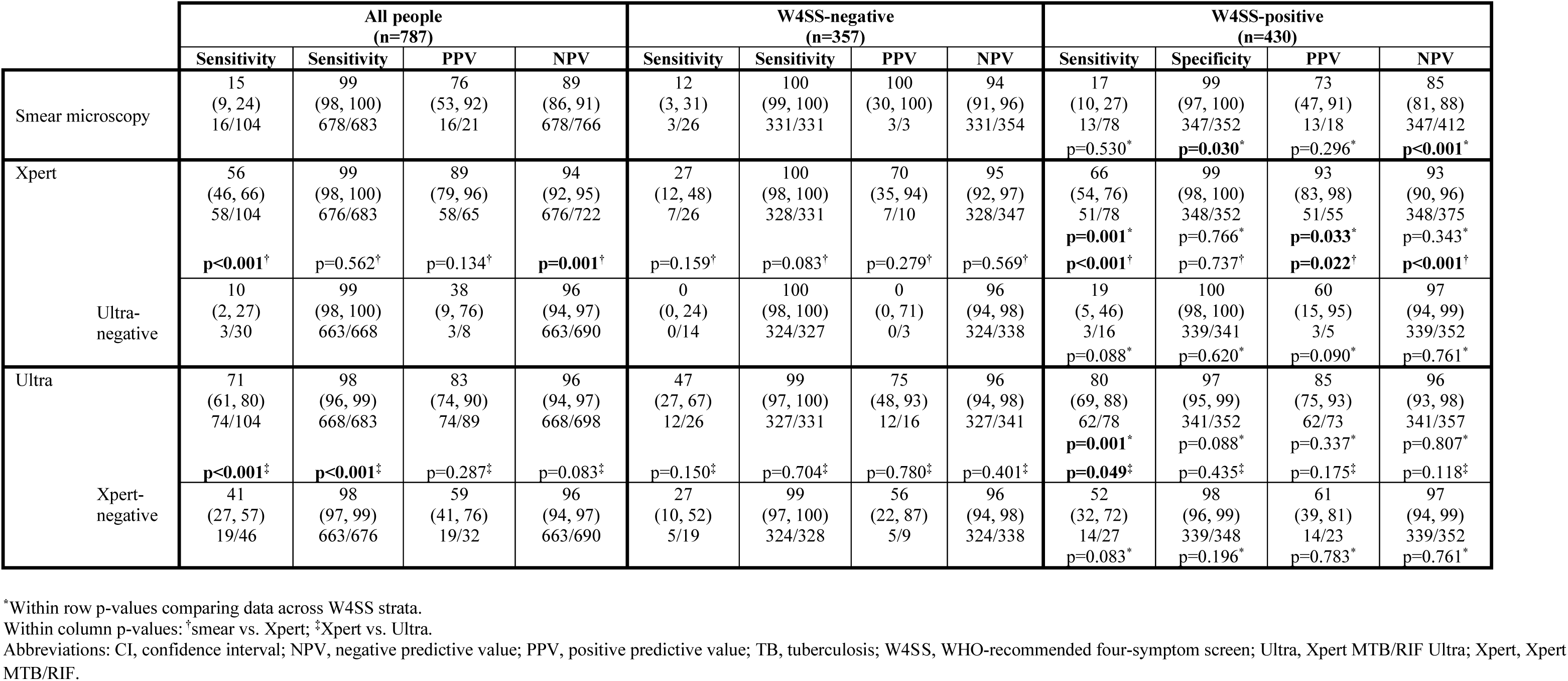
Head-to-head diagnostic accuracy of sputum Xpert and Ultra for TB stratified by W4SS status. Ultra’s sensitivity was higher than Xpert, with Ultra detecting W4SS-negatives missed by Xpert. Ultra specificity decreased compared to Xpert. Sensitivities were higher in W4SS-positives than -negatives. Data are %, 95% CI, and n/N.

#### By CD4 count

Ultra had decreased sensitivity in people with CD4 counts >350 vs. ≤350 cells/µl [42% (19, 68) vs. 77% (66, 86)] and similar specificity [99% (97, 100) vs. 97% (95, 99)] (**Supplementary Table 7**). Within CD4 count strata, Ultra sensitivity was higher than Xpert in people with counts ≤350 cells/µl [77% (66, 86) vs. 60% (48, 71)] and specificity was [97% (95, 99) vs. 99% (98, 100). Similar trends were observed in non-head-to-head data (**Supplementary Table 9**).

#### By W4SS

Ultra’s sensitivity differed from Xpert’s in W4SS-positives [80% (69, 88) vs. 66% (54, 76)] (**Table 4**). Trends in Ultra and Xpert comparative accuracy were similar in smear-negative people and non-head-to-head data (**Supplementary Table 8, 10**). Specificities were similar irrespective of W4SS status.

#### By previous TB

Ultra’s specificity did not differ by previous TB status [97% (91, 99) vs. 98% (97, 99); p=0.50] **(Supplementary Table 11).**

#### Ultra “trace” recategorisation strategies

Reclassifying trace results from positive-to-negative resulted in similar sensitivity [71% (61, 80) vs. 66% (57, 76); Δ-5% (−10, 0); p=0.454] and increased specificity [98% (96,99) vs. 99% (99,100); +Δ1% (1, 3); p=0.011] **(Supplementary Table 12)**. Trace exclusion had a similar effect. Conclusions were unchanged when stratified by previous TB status.

### Urine Ultra and LF-LAM

#### Ultra non-actionable results

Ultra results on concentrated urine were frequently non-actionable [16% (118/732)]. When an unconcentrated aliquot was tested, 99% (117/118) of non-actionables resolved and 11% (13/117) were positive (**Supplementary text, Supplementary Figure 3A**).

#### Sensitivity and specificity

Of the concentrated urine Ultra-positives, 73% (23/33) had a corresponding unconcentrated urine test Ultra-positive (**Supplementary text, Supplementary Figure 3B)**. Amongst the 732 PLHIV in the confirmatory urine test analysis, Ultra and LF-LAM had low sensitivities [25% (17, 35) vs. 15% (9, 24); p=0.070] and high specificities [99 (98, 100) vs. 99 (98, 100); p=0.248] (**Figure 2C, Supplementary Table 13**).

#### Overlap with other tests

Ultra and LF-LAM overlap was high, with each test infrequently giving the only positive non-sputum result [3% (5/129) and 7% (9/129), respectively] (**Supplementary Figure 2**). Five culture-negative people were urine Ultra-positive and, of these, four had a later sputum-based programmatic diagnosis of TB within a year of recruitment (one had no data).

### Sputum scarcity and sputum and urine test yield

#### Sputum scarcity

Amongst people in whom it was known whether sputum induction was necessary, 31% [49/158 (24, 39)] were sputum-scarce (11 W4SS-negative, 38 W4SS-positive), of which 20% [10/49 (10, 34) were culture-positive (all W4SS-positive); comparable to the culture-positivity rate among people who could expectorate [14% (15/109; 7, 22)] **(Supplementary Table 14)**.

#### Overall yield

17% (152/897; 15, 20) of people had at least one positive sputum (culture, Xpert, Ultra) or urine (Ultra, LF-LAM) result. When limited to people in which each test was available and the absence of a result was not due to human error or stock outs, the proportion any-test-positive was 18% (145/804; 15, 21). Sputum tests had highest yields: 71% (103/145; 63, 78) for culture [34% (36/107; 25, 43) single culture-positive], 61% (89/145; 53, 69) for Ultra and 44% (64/145; 36, 53) for Xpert. Urine tests had lower yields with 30% (43/145; 22, 38) and 17% (25/145; 12, 24) positive by Ultra and LF-LAM, respectively.

#### Yield in sputum-scarce people

Of the 158 people in whom it was known whether expectorated sputum could be produced before induction was done, 126 had all the above tests attempted. 23% (29/126; 16, 21) had ≥1 positive result. Of these, 83% (24/29; 64, 95), 66% (19/29; 46, 82) and 59% (17/29; 39, 76) had sputum (expectorated or induced) positive by culture, Ultra, and Xpert respectively. 34% (10/29; 18, 54) and 24% (7/29; 10, 44) had urine positive by Ultra and LF-LAM, respectively. 33% (42/126; 25, 42) were unable to naturally expectorate and induction was their only source of sputum. If induction was done, yields of the aforementioned sputum tests would decrease to 55% (16/29; 36, 74), 45% (13/29; 26, 64), and 38% (11/29; 21, 58), respectively. The aforementioned urine tests’ yields were 17% (5/29; 6, 36) and 10% (3/29; 2, 27), respectively. Comparisons of tests did not show yield and sensitivity and specificity differences when results from expectorated or induced sputum were compared **(Supplementary Table 15)**.

## Discussion

Our key findings are: 1) CRP testing is feasible at POC and superior to W4SS for triaging ART-initiators, reducing unnecessary referrals, improving NNT, and approaching but not capable of meeting the WHO TPP minimum sensitivity and specificity benchmarks, 2) triage algorithms combining W4SS and CRP approach the WHO-recommended optimal specificity target but result in more missed TB than CRP alone, 3) sputum Ultra is more sensitive than Xpert for culture-confirmed TB which is common, both in W4SS-negative and sputum-scarce people, 4) offering sputum induction enhances yield (beyond that possible using urine tests), and 5) urine Ultra and LF-LAM have similar performance and urine Ultra is hampered by high non-actionable result rates. These data inform triage and confirmatory testing strategies, including specimen acquisition, in ART-initiators.

PLHIV should be screened for TB, a process most efficient at ART initiation when patients are immunosuppressed and have relatively high pre-test probability of TB. ART initiators are already within HIV treatment cascades, representing a captured population in a setting in which that we now show CRP measurement at POC is highly feasible and a better alternative to symptoms, significantly reducing unnecessary onward referrals (from 452 to 314 people per 1000 using CRP_10_, translating into a NNT reduction of ∼7 to ∼5). In line with recent WHO guidance^16^, our data also informs the use of CRP at this threshold of ≥10 mg/l, with lower thresholds potentially negating benefits CRP offers over W4SS. Importantly, and in contrast to recent community-based evaluations of CRP^25^, we show little added benefit of combining W4SS with CRP in ART initiators, unless large TB case detection reductions are acceptable, and that haemoglobin should not be considered further due to low sensitivity.

Ultra, despite being WHO-recommended, is unevaluated in ART-initiators without syndromic preselection. We now provide the first data to support its use in this diagnostically-challenging population versus the previous generation Xpert, where sensitivity was higher (71% vs. 56%), including in people who did not meet the W4SS and in people with advanced HIV (CD4 counts <350 cells/µl). This supports Ultra’s use as part of recent efforts to move beyond symptom criteria in high burden settings^26^. Encouragingly, we did not observe dramatically reduced Ultra specificity in contrast to previous research in our setting, however, such research in self-reporting people with presumptive TB, who have a higher rate of previous TB that unselected ART initiators^23^.

Although non-sputum tests are a priority for TB control, there are, unfortunately, unlikely to be alternatives available in the short-term for bacteriological confirmatory testing^1^. Furthermore, countries are heavily invested in sputum-based testing infrastructure and may wish to consider induction, which our data support the use of: 31% required induction to make at least one sputum and without induction the proportion of people with a positive confirmatory test result detected by Ultra would reduce from 65% to 45%. We therefore recommend that programmes consider making sputum induction available at ART-initiation sites, which is currently not standard-of-care in most high burden settings including South Africa. This is especially important given the relatively poor performance of urine tests (although LF-LAM has low yield, it should, due to its low cost, POC nature and, unlike urine Ultra, absence of non-actionable results, still be available in line with current guidance^27^). We also suggest that the utility of sputum induction as a diagnostic intervention, as well as associated implementation challenges like biosafety, requires further evaluation as, based on our data, induction improved case detection more than a new diagnostic test (i.e., Ultra vs. Xpert).

This study has strengths and limitations. We included people irrespective of symptoms, did CRP at POC, utilised a two-sputa culture reference standard, and offered all people sputum induction, however, due to database error, could only confidently establish if induction was necessary in a consecutive subset. Such data are generally limited and our study is still one of only a few to describe sputum production needs in detail in PLHIV^18, 28^, which informs on the potential impact of induction facilities and non-sputum tests. Other limitations include haemoglobin was done in a programmatically-selected subset, which may bias accuracy estimates. We did not analyse the cost, cost-effectiveness, or affordability and these require study. Furthermore, shifting away from decades of entrenched W4SS-based triage in this population will present significant implementation challenges. Barriers to POC CRP implementation require identification, including detailing its impact on patient important outcomes in clinical trials.

In summary, POC CRP is an alternative TB triage tool that has improved specificity compared to self-reported W4SS in PLHIV initiating ART that reduces unnecessary testing. Ultra has improved sensitivity compared to Xpert and detects W4SS-negative TB, and Ultra’s yield is significantly enhanced through sputum induction provision. These tools should be pursued for implementation in this key risk group, together with further evaluations in different populations and settings.

## Data Availability

All data produced in the present study are available upon reasonable request to the authors

## Acknowledgements

The authors thank the participants, Ms Fikiswa Seti, Sr Charmaine Van der Walt, Dr Kim Stanley, and A/Prof Gian van der Spuy. The authors thank the Department of Health, Western Cape Data Warehouse team for providing TB testing and treatment data for patients after the study visit.

## Contributions of authors

GT conceived and designed the study. GT, RW, and PVH acquired funding. Clinical staff collected sample and curated clinical data. Laboratory analysis was done by GN, LR, HT, DM, and ZP. BWPR, GN, HM and GT did analyses and wrote the first draft. All authors critiqued analyses and revised the manuscript.

