## Supplement for "Point-of-care C-reactive protein and Xpert MTB/RIF Ultra for tuberculosis screening and diagnosis in unselected antiretroviral therapy initiators: a prospective diagnostic accuracy study"

### Methods

Definitions

*Time-to-positivity*

Days for a culture to become *Mycobacterium tuberculosis* complex (*Mtb*) positive in the absence of contamination. If both cultures were positive, the shorter time-to-positivity (TTP) was used.

*Ultra "trace reclassified" and "trace excluded"*

"Trace reclassified" treats Ultra-positives with a trace result as Ultra-negative. "Trace excluded" excludes participants with trace results from a particular analysis.

*Non-actionable result*

Any result that is not positive or negative and hence does not provide potentially actionable information to aid clinical decision making. Actionable results are therefore positive or negative or, for a drug susceptibility test, resistant or susceptible.

Specimen processing and diagnostic testing

*C-reactive protein*

iChromaII has a detection range of 2.5-300 mg/l[1]. Values reported by the machine as <2.5 mg/l (n=133) were classified as index test negative. For continuous comparisons involving the n=113, a midpoint value of 1.25 mg/l was arbitrarily assigned. C-reactive protein (CRP) testing was not done for the first 55 participants (no blood collected). A further 150 participants were not tested prospectively due to reagent shortages, but plasma stored at -80°C was tested retrospectively.

*Sputum Xpert and Ultra*

700 µl of one sputum sediment remaining post-culture inoculation was, following storage at -20°C for a maximum of two weeks, selected for Xpert[2] [sediments ≤0.7ml were made up to volume with phosphate buffered saline (Sigma Aldrich, Modderfontein, South Africa)]. Ultra

(version 4) and Xpert (version 2) were done per manufacturer specifications[3,4] (Cepheid, United States) and, if either result was non-actionable (neither positive or negative; see **Definitions** in **Supplement**), specimen-sample reagent mixes were re-tested (if insufficient volume existed the initial result was reported).

*Speciation and drug susceptibility testing*

MTBDR*plus* (Hain Lifesciences, Nehren, Germany) was done on Auramine-O acid fast-positive culture growth (when two cultures were positive, the one with the shortest TTP was tested) for *Mtb* and rifampicin- and isoniazid-susceptibility detection. MTBDR*plus* on the culture isolate served as a reference standard for rifampicin resistance.

*Urine testing*

For Ultra, most urine was tested prospectively [98% (716/732)]. 20 ml was tested or, if less was available, the entire volume used [99% (729/732) of participants had at least 20 ml available]. After centrifugation (4000 rpm, 10 min), supernatant was decanted until a ~700 µl pellet-supernatant mixture was retained prior to resuspension and testing[4]. If the concentrated result was non-actionable or positive, Ultra was repeated on 700 µl unconcentrated urine. For Determine TB LAM (LF-LAM; Abbott, South Africa), unconcentrated urine (60 µl) was tested as recommended[5]. 96% (701/732) participants were tested prospectively and the remainder tested retrospectively due to stock shortages.

### Results

Rifampicin resistance

*Ultra*

Amongst Ultra-positives in the head-to-head sputum comparison with Xpert, 3% (3/89), 80% (71/89) and 17% (15/89) were Ultra rifampicin-resistant, -susceptible, and -indeterminate (all indeterminates trace), respectively. All three Ultra rifampicin-resistant people were MTBDR*plus* rifampicin-resistant and one MTBDR*plus* rifampicin-resistant person was Ultra rifampicin-susceptible [Ultra resistance sensitivity hence 75% (3/4)]. Of the Ultra rifampicin-susceptibles, 6% (4/71) were culture-negative. Ultra's specificity for rifampicin resistance was therefore 99% (66/67). Of the Ultra rifampicin-indeterminate people, 73% (11/15) were culture-negative and the four culture-positives were all MTBDR*plus* rifampicin-susceptible.

*Xpert*

Amongst Xpert-positives in the head-to-head sputum comparison with Ultra, 9% (6/65) were Xpert rifampicin-resistant, 88% (57/65) were Xpert rifampicin-susceptible and 3% (2/65) were rifampicin indeterminate. Of the Xpert resistant cases, 83% (5/6) were culture-positive and MTBDR*plus*-resistant [Xpert sensitivity for rifampicin resistance 100% (5/5)]. Ultra detected 50% (3/6) of these people as rifampicin resistant (of the others, two were Ultra-negative and the other Ultra rifampicin-susceptible). Of the Xpert rifampicin-susceptibles, 7% (4/57) MTBDR*plus* were culture-negative and, of the remaining culture-positives, 100% (53/53) were MTBDR*plus*-susceptible [Xpert specificity for resistance therefore 100% (53/53)]. Two Xpert rifampicin-indeterminates occurred and both were culture-negative.

Urine testing

*Sensitivity and specificity*

No differences were observed between tests when stratified by W4SS-positive or -negatives nor between tests after W4SS stratification. Similar trends occurred for non-head-to-head data.

*Concentrated vs. unconcentrated Ultra*

Concentrated Ultra initially non-actionable: The unconcentrated urine Ultra-positivity rate in people with a non-actionable result from concentrated urine was more than that in people whose concentrated urine was positive [11% (13/118) vs. 5% (33/732); p=0.001], indicating that, in the event of non-actionable concentrated urine Ultra, that participant has increased odds of Ultra-positivity on a corresponding unconcentrated urine **(Supplementary Figure 3A)**.

Concentrated Ultra initially positive: Of the 772 concentrated urines that received Ultra. 4% (33/772) had a positive result [27% (9/33) negative when unconcentrated urine was used, indicating concentration increases yield] and 15% (118/772) were non-actionable (**Supplementary Figure 3B**, remainder of unconcentrated results negative). 100% (33/33) of the concentrated Ultra-positives received unconcentrated Ultra: 97% (32/33) were actionable (one non-actionable) and 72% (23/32) were positive [83% (19/23) rifampicin-susceptible, 4% (1/23) rifampicin-resistant, 9% (2/23) rifampicin-indeterminate, 4% (1/23) rifampicin "no result"].

**Supplementary Figure 1.** ROC curves of haemoglobin among all participants (**A**) and after W4SS (**B, C**) or CD4 count stratification (**D, E**). AUCROCs showed a trend towards being reduced in W4SS-negatives (vs. -positives) and in people with a CD4 count >350 cells/µl (vs. ≤350 cells/µl), but no significant differences were detected. Minimal and optimal sensitivity and specificity for the triage test TPP are represented by single and double ticks, respectively. Grey lines are 95% CIs.

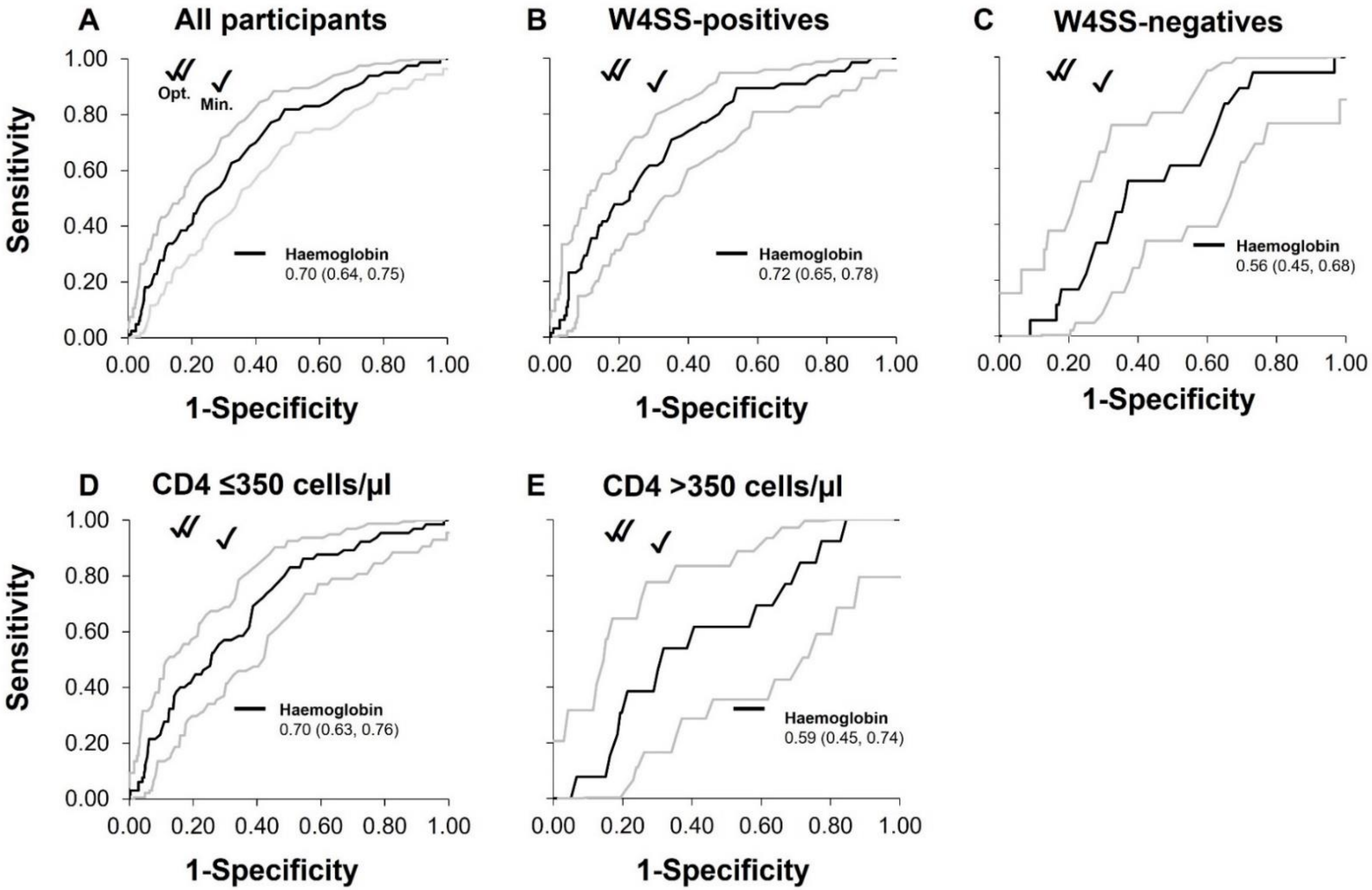

Abbreviations: AUCROC, area under the receiver operator characteristic curve; CI, confidence interval; ROC, receiver operator characteristic; TPP, target product profile; W4SS, World Health Organization-recommended four-symptom screen; WHO, World Health Organization.

**Supplementary Figure 2.** Euler diagrams showing overlap between sputum- and non-sputum tests against composite (**A-C**) and individual (**D-F**) culture results. Numbers represent people positive for each test on sputum (first column), urine (with the sputum reference standard; second column), or sputum and urine (third column). (**A**) Sputum Ultra detected most culture-positives missed by Xpert and, in 10% (13/129) of people with a positive sputum test result, was the only positive result. (**B**) Urine Ultra and LF-LAM had poor yield and missed many cases but detected people missed by other tests (including culture). (**C**) When comparing tests on sputum and urine, each test exclusively detected some people missed by other tests. **D-F** show many people were positive only by a single culture, and a two-culture reference standard's importance. Note people require a positive or negative result to be included here, which is the why total n varies slightly across panels and versus yield calculations (**Results**).

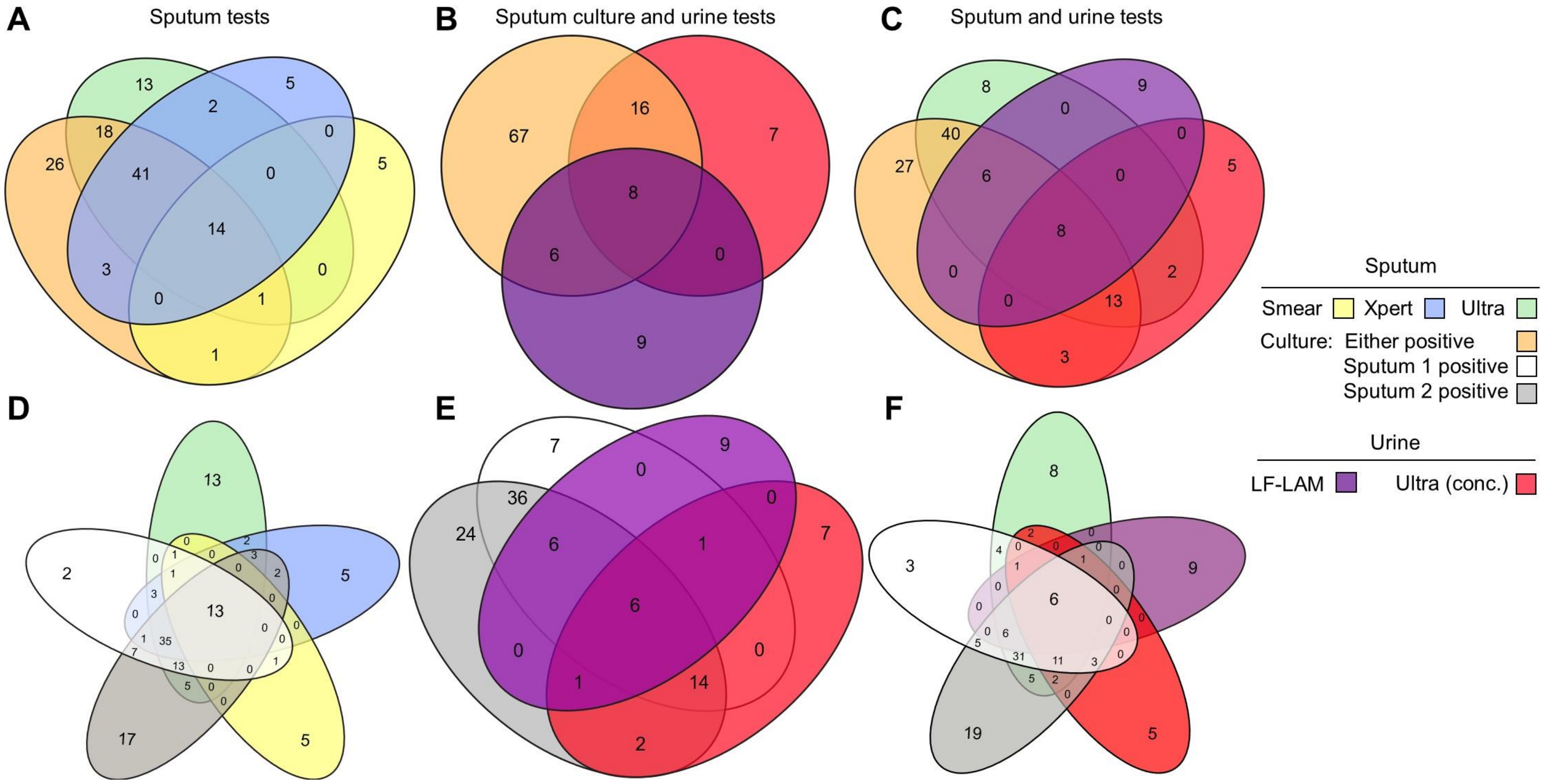

Abbreviations: conc., concentrated. LF-LAM, Determine TB LAM Ag test; MGIT, Mycobacteria Growth Indicator Tube; Ultra, Xpert MTB/RIF Ultra; Xpert, Xpert MTB/RIF.

**Supplementary Figure 3.** Unconcentrated urine Ultra results in participants who had, on a paired concentrated urine, a (**A**) non-actionable or (**B**) positive result. 27% (9/33) of concentrated Ultra-positive urines were negative when unconcentrated urine was tested, indicating concentration increases yield. Of the concentrated urine non-actionable results, 99% (117/118) resolved when unconcentrated urine was tested, and 11% (13/117) of these were Ultra-positive upon retesting (for two people, this unconcentrated Ultra result was the only positive confirmatory test result). The unconcentrated urine Ultra-positivity rate in people with a non-actionable result from concentrated urine was more than double that in people whose concentrated urine was positive [11% (13/118) vs. 5% (33/732); p=0.001], indicating that, in the event of non-actionable concentrated urine Ultra, the participant has increased odds of Ultra-positivity on a corresponding unconcentrated urine. Abbreviations: RIF, rifampicin; Ultra, Xpert MTB/RIF Ultra.

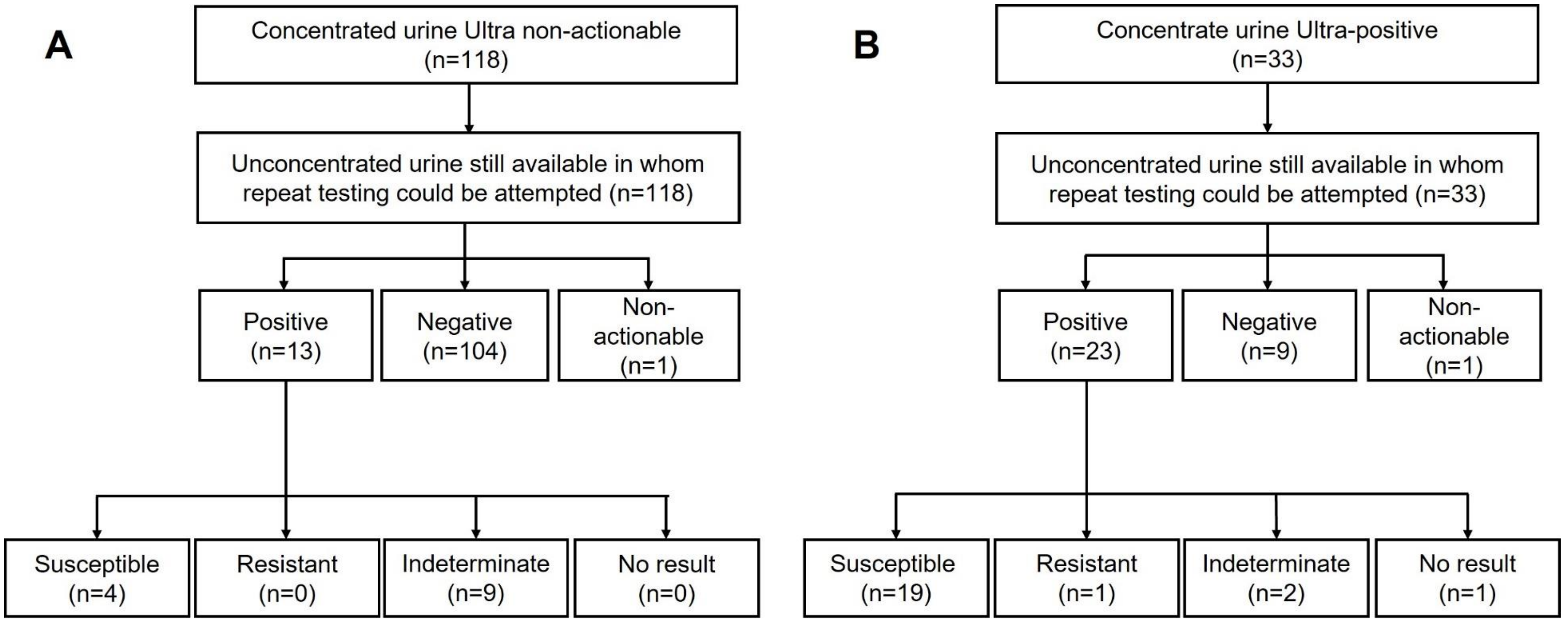

**Supplementary Table 1.** Head-to-head diagnostic accuracy of cough and CRP stratified by W4SS status, shown overall and in smear-negative people only. CRP had decreased sensitivity in W4SS-negatives vs. -positives but higher specificity. When comparing CRP thresholds within people of the same W4SS status, ≥10 mg/l had higher specificity than ≥5 mg/l. Trends were similar in smear-negative people. The "All" column (for the "irrespective of smear status" section) is the same as in **Table 2**. Data are %, 95% CI, and n/N.

| | **All (n=800)** | | | | **W4SS-positives (n=442)** | | | | **W4SS-negatives (n=358)** | | | |
|---|---|---|---|---|---|---|---|---|---|---|---|---|
| | **Sensitivity** | **Specificity** | **PPV** | **NPV** | **Sensitivity** | **Specificity** | **PPV** | **NPV** | **Sensitivity** | **Specificity** | **PPV** | **NPV** |
| Irrespective of smear status | | | | | | | | | | | | |
| Cough (any duration) | 53<br>(43, 63)<br>55/104 | 73<br>(70, 76)<br>506/696 | 22<br>(18, 29)<br>55/245 | 91<br>(89, 94)<br>506/555 | 69<br>(58, 79)<br>55/80 | 48<br>(43, 53)<br>172/362 | 22<br>(18, 29)<br>55/245 | 87<br>(82, 92)<br>172/197 | 0<br>(0, 15)<br>0/24<br>**p<0.001**[**] | 100<br>(99, 100)<br>334/334<br>**p<0.001**[**] | Non-calculable<br>- | 93<br>(91, 96)<br>334/358<br>**p=0.017**[**] |
| ≥2 weeks | 42<br>(33, 53)<br>44/104<br>p=0.127[*] | 83<br>(81, 86)<br>579/696<br>**p<0.001**[*] | 27<br>(21, 35)<br>44/161<br>p=0.263[*] | 91<br>(89, 93)<br>579/639<br>p=0.737[*] | 55<br>(44, 67)<br>44/80<br>p=0.073[*] | 68<br>(63, 73)<br>245/362<br>**p<0.001**[*] | 27<br>(21, 35)<br>44/161<br>p=0.263[*] | 87<br>(83, 91)<br>245/281<br>p=0.969[*] | 0<br>(0, 15)<br>0/24<br>**p<0.001**[**]<br>- | 100<br>(99, 100)<br>334/334<br>**p<0.001**[**]<br>- | Non-calculable<br>-<br>- | 93<br>(91, 96)<br>334/358<br>**p=0.009**[**]<br>p>0.999[*] |
| CRP<br>≥5 mg/l | 86<br>(78, 92)<br>89/104<br>**p<0.001**[†] | 49<br>(45, 53)<br>339/696<br>**p<0.001**[†] | 20<br>(17, 24)<br>89/446<br>p=0.440[†] | 96<br>(94, 98)<br>339/354<br>**p=0.008**[†] | 94<br>(87, 98)<br>75/80<br>**p<0.001**[†] | 40<br>(35, 45)<br>143/362<br>**p=0.030**[†] | 26<br>(21, 31)<br>75/294<br>p=0.408[†] | 97<br>(93, 99)<br>143/148<br>p=0.969[†] | 58<br>(37, 78)<br>14/24<br>**p<0.001**[**]<br>**p<0.001**[†] | 59<br>(54, 65)<br>196/334<br>**p<0.001**[**]<br>**p<0.001**[†] | 9<br>(6, 15)<br>14/152<br>**p<0.001**[**]<br>- | 95<br>(92, 98)<br>196/206<br>p=0.497[**]<br>p=0.374[†] |
| ≥10 mg/l | 77<br>(68, 85)<br>80/104<br>**p<0.001**[†]<br>p=0.394[‡] | 64<br>(61, 68)<br>445/696<br>**p<0.001**[†]<br>**p<0.001**[‡] | 24<br>(20, 30)<br>80/331<br>p=0.630[†]<br>p=0.159[‡] | 95<br>(93, 97)<br>445/469<br>**p=0.021**[†]<br>p=0.556[‡] | 90<br>(82, 96)<br>72/80<br>**p=0.001**[†]<br>p=0.385[‡] | 54<br>(49, 59)<br>194/362<br>p=0.102[†]<br>**p<0.001**[‡] | 30<br>(25, 37)<br>72/240<br>p=0.059[†]<br>p=0.248[‡] | 96<br>(93, 99)<br>194/202<br>**p=0.002**[†]<br>p=0.776[‡] | 33<br>(16, 56)<br>8/24<br>**p<0.001**[**]<br>**p=0.002**[†]<br>p=0.082[‡] | 75<br>(71, 80)<br>251/334<br>**p<0.001**[**]<br>**p<0.001**[†]<br>**p<0.001**[‡] | 9<br>(4, 17)<br>8/91<br>**p<0.001**[**]<br>-<br>p=0.912[‡] | 94<br>(91, 97)<br>251/267<br>p=0.323[**]<br>p=0.719[†]<br>p=0.590[‡] |
| Smear-negatives only | | | | | | | | | | | | |
| Cough (any duration) | 49<br>(39, 60)<br>43/88 | 73<br>(70, 77)<br>504/691 | 19<br>(14, 25)<br>43/230 | 92<br>(90, 95)<br>504/549 | 64<br>(52, 76)<br>43/67 | 4<br>(43, 53)<br>170/357 | 19<br>(14, 25)<br>43/230 | 88<br>(83, 92)<br>170/194 | 0<br>(0, 17)<br>0/21<br>**p<0.001**[**] | 100<br>(99, 100)<br>334/334<br>**p<0.001**[**] | Non-calculable<br>- | 94<br>(92, 97)<br>334/355<br>**p=0.008**[**] |
| ≥2 weeks | 38<br>(28, 49)<br>33/88<br>p=0.128[*] | 84<br>(81, 87)<br>576/691<br>**p<0.001**[*] | 22<br>(16, 30)<br>33/148<br>p=0.394[*] | 91<br>(89, 94)<br>576/631<br>p=0.749[*] | 49<br>(37, 62)<br>33/67<br>p=0.081[*] | 68<br>(63, 73)<br>242/357<br>**p<0.001**[*] | 22<br>(16, 30)<br>33/148<br>p=0.394[*] | 88<br>(84, 92)<br>242/276<br>p=0.986[*] | 0<br>(0, 17)<br>0/21<br>**p<0.001**[**]<br>- | 100<br>(99, 100)<br>334/334<br>**p<0.001**[**]<br>- | Non-calculable<br>-<br>- | 94<br>(92, 97)<br>334/355<br>**p=0.005**[**]<br>p>0.999[*] |
| CRP<br>≥5 mg/l | 84<br>(75, 92)<br>74/88<br>**p<0.001**[†] | 49<br>(46, 54)<br>338/691<br>**p<0.001**[†] | 17<br>(14, 22)<br>74/427<br>p=0.663[†] | 96<br>(94, 98)<br>338/352<br>**p=0.012**[†] | 93<br>(84, 98)<br>62/67<br>**p<0.001**[†] | 40<br>(35, 46)<br>142/357<br>**p<0.001**[†] | 22<br>(18, 28)<br>62/277<br>p=0.253[†] | 97<br>(93, 99)<br>142/147<br>**p=0.003**[†] | 57<br>(35, 79)<br>12/21<br>**p<0.001**[**]<br>**p<0.001**[†] | 59<br>(54, 65)<br>196/334<br>**p<0.001**[**]<br>**p<0.001**[†] | 8<br>(5, 14)<br>12/150<br>**p<0.001**[**]<br>- | 96<br>(92, 98)<br>196/205<br>p=0.640[**]<br>p=0.440[†] |

| | | | | | | | | | | | | |
|---|---|---|---|---|---|---|---|---|---|---|---|---|
| ≥10 mg/l | 74<br>(64, 83)<br>65/88<br><br>**p=0.001†**<br>p=0.096‡ | 64<br>(61, 68)<br>444/691<br><br>**p=0.001†**<br>**p<0.001‡** | 21<br>(17, 26)<br>65/312<br><br>p=0.538†<br>p=0.229‡ | 95<br>(93, 97)<br>444/467<br><br>**p=0.038†**<br>p=0.518‡ | 88<br>(78, 95)<br>59/67<br><br>**p=0.001†**<br>p=0.381‡ | 54<br>(49, 60)<br>193/357<br><br>p=0.085†<br>**p<0.001‡** | 26<br>(21, 33)<br>59/223<br><br>**p=0.048†**<br>p=0.290‡ | 96<br>(93, 99)<br>193/201<br><br>**p=0.002†**<br>p=0.778‡ | 29<br>(12, 53)<br>6/21<br>**p<0.001****<br>**p=0.008†**<br>p=0.061‡ | 75<br>(71, 80)<br>251/334<br>**p<0.001****<br>**p<0.001†**<br>**p<0.001‡** | 7<br>(3, 15)<br>6/89<br>**p<0.001****<br>-<br>p=0.721‡ | 94<br>(91, 97)<br>251/266<br>p=0.412**<br>p=0.884†<br>p=0.514‡ |

Within row p-values: **W4SS-positive vs. -negative.
Within column p-values: *Any vs. ≥2-week cough; †Cough (any) vs. CRP; ‡CRP (≥5 vs ≥10 mg/l).
Abbreviations: CRP, C-reactive protein; CI, confidence interval; NPV, negative predictive value; PPV, positive predictive value; W4SS, WHO-recommended four-symptom screen.

**Supplementary Table 2**. Head-to-head diagnostic accuracy of W4SS and CRP stratified by CD4 count in smear-negative people. Trends were like those in all people irrespective of smear status (**Table 2**). Data are %, 95% CI, and n/N.

| | **All (n=735)** | | | | **CD4 count >350 cells/µl (n=309)** | | | | **CD4 count ≤350 cells/µl (n=426)** | | | |
|---|---|---|---|---|---|---|---|---|---|---|---|---|
| | **Sensitivity** | **Specificity** | **PPV** | **NPV** | **Sensitivity** | **Specificity** | **PPV** | **NPV** | **Sensitivity** | **Specificity** | **PPV** | **NPV** |
| Cough (any duration) | 49<br>(39, 60)<br>43/88 | 73<br>(70, 77)<br>504/691 | 19<br>(14, 25)<br>43/230 | 92<br>(90, 95)<br>504/549 | 40<br>(17, 68)<br>6/15 | 80<br>(75, 85)<br>235/294 | 9<br>(4, 20)<br>6/65 | 96<br>(94, 99)<br>235/244 | 50<br>(38, 63)<br>33/66<br>p=0.481** | 68<br>(63, 73)<br>243/360<br>**p<0.001**** | 22<br>(16, 30)<br>33/150<br>**p=0.026**** | 88<br>(84, 92)<br>243/276<br>**p=0.001**** |
| ≥2 weeks | 38<br>(28, 49)<br>33/88<br>p=0.128* | 84<br>(81, 87)<br>576/691<br>**p<0.001*** | 22<br>(16, 30)<br>33/148<br>p=0.394* | 91<br>(89, 94)<br>576/631<br>p=0.749* | 27<br>(8, 56)<br>4/15<br>p=0.439* | 88<br>(84, 92)<br>258/294<br>p=0.010* | 10<br>(3, 24)<br>4/40<br>p=0.896* | 50<br>(93, 98)<br>258/269<br>p=0.815* | 39<br>(28, 53)<br>26/66<br>p=0.357**<br>p=0.220* | 81<br>(77, 85)<br>291/360<br>**p=0.017****<br>**p<0.001*** | 27<br>(19, 38)<br>26/95<br>**p=0.027****<br>p=0.338* | 88<br>(84, 92)<br>291/331<br>**p=0.001****<br>p=0.962* |
| W4SS | 76<br>(66, 85)<br>67/88 | 48<br>(46, 53)<br>334/691 | 16<br>(13, 20)<br>67/424 | 94<br>(92, 97)<br>334/355 | 47<br>(22, 74)<br>7/15 | 54<br>(49, 61)<br>160/294 | 5<br>(3, 10)<br>7/141 | 95<br>(91, 98)<br>160/168 | 82<br>(71, 91)<br>54/66<br>**p=0.004**** | 45<br>(40, 51)<br>161/360<br>**p=0.014**** | 21<br>(17, 27)<br>54/253<br>**p<0.001**** | 93<br>(89, 97)<br>161/173<br>p=0.393** |
| CRP<br>≥5 mg/l | 84<br>(75, 92)<br>74/88<br>p=0.186† | 49<br>(46, 54)<br>338/691<br>p=0.830† | 17<br>(14, 22)<br>74/427<br>p=0.549† | 96<br>(94, 98)<br>338/352<br>p=0.235† | 71<br>(45, 90)<br>10/15<br>p=0.269† | 60<br>(55, 66)<br>176/294<br>p=0.182† | 9<br>(5, 16)<br>10/128<br>p=0.338† | 97<br>(94, 100)<br>176/181<br>p=0.324† | 88<br>(78, 95)<br>58/66<br>**p=0.043****<br>p=0.332† | 41<br>(36, 47)<br>147/360<br>**p=0.001****<br>p=0.292† | 21<br>(17, 27)<br>58/271<br>**p=0.001****<br>p=0.987† | 95<br>(91, 98)<br>147/155<br>p=0.256**<br>p=0.504† |
| ≥10 mg/l | 74<br>(64, 83)<br>65/88<br>p=0.728†<br>p=0.096‡ | 64<br>(61, 68)<br>444/691<br>**p<0.001†**<br>**p<0.001‡** | 21<br>(17, 26)<br>65/312<br>p=0.079†<br>p=0.229‡ | 95<br>(93, 97)<br>444/467<br>p=0.532†<br>p=0.518‡ | 33<br>(12, 62)<br>5/15<br>p=0.456†<br>p=0.068‡ | 72<br>(67, 77)<br>211/294<br>**p<0.001†**<br>**p=0.002‡** | 6<br>(2, 13)<br>5/88<br>p=0.813†<br>p=0.545‡ | 95<br>(92, 98)<br>211/221<br>p=0.912†<br>p=0.354‡ | 82<br>(71, 91)<br>54/66<br>**p<0.001****<br>p>0.999†<br>p=0.332‡ | 59<br>(54, 65)<br>213/360<br>**p=0.001****<br>**p<0.001†**<br>**p<0.001‡** | 27<br>(21, 34)<br>54/201<br>**p<0.001****<br>p=0.170†<br>p=0.168‡ | 95<br>(91, 98)<br>213/225<br>p=0.693**<br>p=0.505†<br>p=0.941‡ |

Within row p-values: **CD4 count >350 vs. ≤350 cells/µl.
Within column p-values: *Any vs. ≥2-week cough; †W4SS vs. CRP; ‡CRP (≥5 vs. ≥10 mg/l).
Missing data: CD4 within three months (n=44).
Abbreviations: CRP, C-reactive protein; CI, confidence interval; NPV, negative predictive value; PPV, positive predictive value; W4SS, WHO-recommended four-symptom screen.

**Supplementary Table 3.** Non-head-to-head diagnostic accuracy of cough and CRP stratified by W4SS status. Findings in this table are like head-to-head data in **Supplementary Table 2**. Data are %, 95% CI, and n/N.

| | **All** | | | | **W4SS-positives** | | | | **W4SS-negatives** | | | |
|---|---|---|---|---|---|---|---|---|---|---|---|---|
| | **Sensitivity** | **Specificity** | **PPV** | **NPV** | **Sensitivity** | **Specificity** | **PPV** | **NPV** | **Sensitivity** | **Specificity** | **PPV** | **NPV** |
| Irrespective of smear status | | | | | | | | | | | | |
| Cough (any duration) | 52<br>(43, 63)<br>56/107 | 73<br>(70, 76)<br>544/748 | 22<br>17, 28)<br>56/260 | 91<br>(89, 94)<br>544/595 | 65<br>(53, 76)<br>44/68 | 47<br>(43, 53)<br>180/381 | 18<br>(14, 24)<br>44/245 | 88<br>(84, 93)<br>180/204 | 0<br>(0, 15)<br>0/23<br>**p<0.001**$^{**}$ | 100<br>(99, 100)<br>362/362<br>**p<0.001**$^{**}$ | Non-calculable<br>- | 94<br>(92, 97)<br>362/385<br>**p=0.014**$^{**}$ |
| ≥2 weeks | 42<br>(33, 52)<br>45/107<br>p=0.132$^{*}$ | 83<br>(81, 86)<br>621/748<br>**p<0.001**$^{*}$ | 26<br>(20, 34)<br>45/172<br>p=0.266$^{*}$ | 91<br>(89, 93)<br>621/683<br>p=0.751$^{*}$ | 50<br>(38, 63)<br>34/68<br>p=0.083$^{*}$ | 67<br>(63, 72)<br>256/381<br>**p<0.001**$^{*}$ | 21<br>(16, 29)<br>34/159<br>p=0.394$^{*}$ | 88<br>(85, 92)<br>256/290<br>p=0.989$^{*}$ | 0<br>(0, 15)<br>0/23<br>**p<0.001**$^{**}$<br>- | 100<br>(99, 100)<br>362/362<br>**p<0.001**$^{**}$<br>- | Non-calculable<br>-<br>- | 94<br>(92, 97)<br>362/385<br>p=0.008$^{**}$<br>- |
| CRP<br>≥5 mg/l | 86<br>(78, 92)<br>89/104<br>**p<0.001**$^{\dagger}$ | 49<br>(45, 53)<br>339/696<br>**p<0.001**$^{\dagger}$ | 20<br>(17, 24)<br>89/446<br>p=0.615$^{\dagger}$ | 96<br>(94, 98)<br>339/354<br>**p=0.011**$^{\dagger}$ | 93<br>(84, 98)<br>62/67<br>**p<0.001**$^{\dagger}$ | 40<br>(35, 46)<br>142/357<br>**p=0.041**$^{\dagger}$ | 22<br>(18, 28)<br>62/277<br>p=0.210$^{\dagger}$ | 97<br>(93, 99)<br>142/147<br>**p=0.005**$^{\dagger}$ | 57<br>(35, 79)<br>12/21<br>**p<0.001**$^{**}$<br>**p<0.001**$^{\dagger}$ | 59<br>(54, 65)<br>196/334<br>**p<0.001**$^{**}$<br>**p<0.001**$^{\dagger}$ | 8<br>(5, 14)<br>12/150<br>**p<0.001**$^{**}$<br>- | 96<br>(92, 98)<br>196/205<br>p=0.640$^{**}$<br>p=0.419$^{\dagger}$ |
| ≥10 mg/l | 77<br>(68, 85)<br>80/104<br>**p<0.001**$^{\dagger}$ | 64<br>(61, 68)<br>445/696<br>**p<0.001**$^{\dagger}$ | 24<br>(20, 30)<br>80/331<br>p=0.451$^{\dagger}$ | 95<br>(93, 97)<br>445/469<br>**p=0.029**$^{\dagger}$ | 88<br>(78, 95)<br>59/67<br>**p=0.001**$^{\dagger}$ | 54<br>(49, 60)<br>193/357<br>p=0.064$^{\dagger}$ | 26<br>(21, 33)<br>59/223<br>**p=0.027**$^{\dagger}$ | 96<br>(93, 99)<br>193/201<br>**p=0.004**$^{\dagger}$ | 29<br>(12, 53)<br>6/21<br>**p<0.001**$^{**}$<br>**p=0.007**$^{\dagger}$ | 75<br>(71, 80)<br>251/334<br>**p<0.001**$^{**}$<br>**p<0.001**$^{\dagger}$ | 7<br>(3, 15)<br>6/89<br>**p<0.001**$^{**}$<br>- | 94<br>(91, 97)<br>251/266<br>p=0.412$^{**}$<br>p=0.858$^{\dagger}$ |
| Smear-negatives only | | | | | | | | | | | | |
| Cough (any duration) | 48<br>(38, 60)<br>44/91 | 73<br>(70, 77)<br>542/743 | 18<br>(14, 24)<br>44/245 | 92<br>(90, 95)<br>542/589 | 65<br>(53, 76)<br>44/68 | 47<br>(43, 53)<br>180/381 | 18<br>(14, 24)<br>44/245 | 88<br>(84, 93)<br>180/204 | 0<br>(0, 15)<br>0/23<br>**p<0.001**$^{**}$ | 100<br>(99, 100)<br>362/362<br>**p<0.001**$^{**}$ | Non-calculable<br>- | 94<br>(92, 97)<br>362/385<br>**p=0.014**$^{**}$ |
| ≥2 weeks | 37<br>(28, 49)<br>34/91<br>p=0.134$^{*}$ | 83<br>(81, 86)<br>618/743<br>**p<0.001**$^{*}$ | 21<br>(16, 29)<br>34/159<br>p=0.394$^{*}$ | 92<br>(90, 94)<br>618/675<br>p=0.764$^{*}$ | 50<br>(38, 63)<br>34/68<br>p=0.083$^{*}$ | 67<br>(63, 72)<br>256/381<br>**p<0.001**$^{*}$ | 21<br>(16, 29)<br>34/159<br>p=0.394$^{*}$ | 88<br>(85, 92)<br>256/290<br>p=0.989$^{*}$ | 0<br>(0, 15)<br>0/23<br>**p<0.001**$^{**}$<br>- | 100<br>(99, 100)<br>362/362<br>**p<0.001**$^{**}$<br>- | Non-calculable<br>-<br>- | 94<br>(92, 97)<br>362/385<br>**p=0.008**$^{**}$<br>p>0.999$^{*}$ |
| CRP<br>≥5 mg/l | 84<br>(75, 92)<br>74/88<br>**p<0.001**$^{\dagger}$ | 49<br>(46, 53)<br>338/691<br>**p<0.001**$^{\dagger}$ | 17<br>(14, 22)<br>74/427<br>p=0.837$^{\dagger}$ | 96<br>(94, 98)<br>338/352<br>**p=0.016**$^{\dagger}$ | 93<br>(84, 98)<br>62/67<br>**p<0.001**$^{\dagger}$ | 40<br>(35, 46)<br>142/357<br>**p<0.001**$^{\dagger}$ | 22<br>(18, 28)<br>62/277<br>p=0.210$^{\dagger}$ | 97<br>(93, 99)<br>142/147<br>**p=0.005**$^{\dagger}$ | 57<br>(35, 79)<br>12/21<br>**p<0.001**$^{**}$<br>**p<0.001**$^{\dagger}$ | 59<br>(54, 65)<br>196/334<br>**p<0.001**$^{**}$<br>**p<0.001**$^{\dagger}$ | 8<br>(5, 14)<br>12/150<br>**p<0.001**$^{**}$<br>- | 96<br>(92, 98)<br>196/205<br>p=0.640$^{**}$<br>p=0.419$^{\dagger}$ |
| ≥10 mg/l | 74<br>(64, 83)<br>65/88 | 64<br>(61, 68)<br>444/691 | 21<br>(17, 26)<br>65/312 | 95<br>(93, 97)<br>444/467 | 88<br>(78, 95)<br>59/67 | 54<br>(49, 60)<br>193/357 | 26<br>(21, 33)<br>59/223 | 96<br>(93, 99)<br>193/201 | 29<br>(12, 53)<br>6/21<br>**p<0.001**$^{**}$ | 75<br>(71, 80)<br>251/334<br>**p<0.001**$^{**}$ | 7<br>(3, 15)<br>6/89<br>**p<0.001**$^{**}$ | 94<br>(91, 97)<br>251/266<br>p=0.412$^{**}$ |

| | | | | | | | | | | | | |
|---|---|---|---|---|---|---|---|---|---|---|---|---|
| | **p<0.001**[†] | **p<0.001**[†] | p=0.396[†] | **p=0.048**[†] | **p=0.001**[†] | p=0.064[†] | **p=0.027**[†] | **p=0.004**[†] | **p=0.006**[†] | **p<0.001**[†] | - | p=0.858[†] |

Within row p-values: [**]W4SS-positives vs -negatives.
Within column p-values: [*]Any vs. ≥2-week cough; [†]Cough (any) vs. CRP.
Abbreviations: CRP, C-reactive protein; CI, confidence interval; NPV, negative predictive value; PPV, positive predictive value; W4SS, WHO-recommended four-symptom screen.

**Supplementary Table 4.** Non-head-to-head diagnostic accuracy data of potential triage tests ("all" is the same as in **Supplementary Table 2**). Diagnostic accuracy was like the head-to-head analyses done overall (**Table 2**) and in smear-negatives (**Supplementary Table 2**). Data are %, 95% CI, and n/N.

| | **All** | | | | **CD4 count >350 cells/µl** | | | | **CD4 count ≤350 cells/µl** | | | |
|---|---|---|---|---|---|---|---|---|---|---|---|---|
| | **Sensitivity** | **Specificity** | **PPV** | **NPV** | **Sensitivity** | **Specificity** | **PPV** | **NPV** | **Sensitivity** | **Specificity** | **PPV** | **NPV** |
| Irrespective of smear status | | | | | | | | | | | | |
| Cough (any duration) | 52<br>(43, 63)<br>56/107 | 73<br>(70, 76)<br>544/748 | 22<br>17, 28)<br>56/260 | 91<br>(89, 94)<br>544/595 | 39<br>(18, 65)<br>7/18 | 80<br>(75, 84)<br>246/309 | 10<br>(5, 20)<br>7/70 | 96<br>(93, 98)<br>246/257 | 53<br>(42, 65)<br>42/79<br>p=0.274[**] | 68<br>(63, 73)<br>267/395<br>**p=0.001**[**] | 25<br>(19, 32)<br>42/170<br>**p=0.010**[**] | 88<br>(84, 92)<br>267/304<br>**p=0.001**[**] |
| ≥2 weeks | 42<br>(33, 52)<br>45/107<br>p=0.132[*] | 83<br>(81, 86)<br>621/748<br>**p<0.001**[*] | 26<br>(20, 34)<br>45/172<br>p=0.266[*] | 91<br>(89, 93)<br>621/683<br>p=0.751[*] | 28<br>(10, 54)<br>5/18<br>p=0.480[*] | 88<br>(84, 92)<br>271/309<br>**p<0.007**[*] | 12<br>(4, 26)<br>5/43<br>p=0.785[*] | 95<br>(93, 98)<br>271/284<br>p=0.867[*] | 43<br>(32, 55)<br>34/79<br>p=0.233[**]<br>p=0.203[*] | 80<br>(76, 85)<br>317/395<br>**p=0.008**[**]<br>**p<0.001**[*] | 30<br>(23, 40)<br>34/112<br>**p=0.016**[**]<br>p=0.295[*] | 88<br>(84, 91)<br>317/362<br>**p=0.001**[**]<br>p=0.919[*] |
| W4SS | 76<br>(67, 84)<br>81/107 | 48<br>(45, 53)<br>362/748 | 17<br>(15, 22)<br>81/467 | 93<br>(91, 96)<br>362/388 | 50<br>(27, 74)<br>9/18 | 55<br>(50, 61)<br>171/309 | 6<br>(3, 12)<br>9/147 | 95<br>(91, 98)<br>171/180 | 80<br>(70, 88)<br>63/79<br>**p=0.009**[**] | 44<br>(40, 50)<br>174/395<br>**p=0.003**[**] | 22<br>(18, 28)<br>63/284<br>**p<0.001**[**] | 92<br>(87, 96)<br>174/190<br>**p=0.190**[**] |
| CRP<br>≥5mg/l | 86<br>(78, 92)<br>89/104<br>p=0.070[†] | 49<br>(45, 53)<br>339/696<br>p=0.906[†] | 20<br>(17, 24)<br>89/446<br>p=0.311[†] | 96<br>(94, 98)<br>339/354<br>p=0.142[†] | 67<br>(41, 87)<br>12/18<br>p=0.310[†] | 60<br>(55, 66)<br>176/294<br>p=0.261[†] | 9<br>(5, 16)<br>12/130<br>p=0.329[†] | 97<br>(93, 99)<br>176/182<br>p=0.416[†] | 89<br>(81, 96)<br>68/76<br>**p=0.015**[**]<br>p=0.094[†] | 41<br>(36, 46)<br>148/365<br>**p<0.001**[**]<br>p=0.329[†] | 24<br>(20, 30)<br>68/285<br>**p<0.001**[**]<br>p=0.645[†] | 95<br>(91, 98)<br>148/156<br>**p=0.004**[**]<br>p=0.230[†] |
| ≥10mg/l | 77<br>(68, 85)<br>80/104<br>p=0.840[†] | 64<br>(61, 68)<br>445/696<br>**p<0.001**[†] | 24<br>(20, 30)<br>80/331<br>**p=0.018**[†] | 95<br>(93, 97)<br>445/469<br>p=0.325[†] | 39<br>(18, 65)<br>7/18<br>p=0.502[†] | 72<br>(67, 77)<br>211/294<br>**p<0.001**[†] | 8<br>(4, 16)<br>7/90<br>p=0.622[†] | 95<br>(92, 98)<br>211/222<br>p=0.984[†] | 84<br>(75, 92)<br>64/76<br>**p<0.001**[**]<br>p=0.470[†] | 59<br>(54, 64)<br>214/365<br>**p<0.001**[**]<br>**p<0.001**[†] | 30<br>(24, 37)<br>64/215<br>**p<0.001**[**]<br>p=0.054[†] | 95<br>(91, 98)<br>214/226<br>p=0.865[**]<br>p=0.207[†] |
| Smear-negatives | | | | | | | | | | | | |
| Cough (any duration) | 48<br>(38, 60)<br>44/91 | 73<br>(70, 77)<br>542/743 | 18<br>(14, 24)<br>44/245 | 92<br>(90, 95)<br>542/589 | 40<br>(17, 68)<br>6/15 | 80<br>(75, 84)<br>246/309 | 9<br>(4, 18)<br>6/69 | 96<br>(94, 99)<br>246/255 | 49<br>(90, 100)<br>34/69<br>p=0.515[**] | 68<br>(64, 73)<br>265/390<br>**p=0.001**[**] | 21<br>(16, 29)<br>34/159<br>**p=0.021**[**] | 88<br>(85, 92)<br>265/300<br>**p<0.001**[**] |
| ≥2 weeks | 37<br>(28, 49)<br>34/91<br>p<0.134[*] | 83<br>(81, 86)<br>618/743<br>**p<0.001**[*] | 21<br>(16, 29)<br>34/159<br>p=0.394[*] | 92<br>(90, 94)<br>618/675<br>**p<0.001**[*] | 27<br>(8, 56)<br>4/15<br>p=0.439[*] | 88<br>(84, 92)<br>271/309<br>**p=0.007**[*] | 10<br>(3, 23)<br>4/42<br>p=0.883[*] | 96<br>(94, 99)<br>271/282<br>p=0.821[*] | 39<br>(88, 100)<br>27/69<br>p=0.365[**]<br>p=0.230[*] | 81<br>(77, 85)<br>314/390<br>**p=0.011**[**]<br>**p<0.001**[*] | 26<br>(19, 36)<br>27/103<br>**p=0.026**[**]<br>p=0.366[*] | 88<br>(85, 92)<br>314/356<br>**p<0.001**[**]<br>p=0.959[*] |
| W4SS | 75<br>(65, 84)<br>68/91 | 49<br>(46, 53)<br>362/743 | 15<br>(12, 19)<br>68/449 | 94<br>(92, 97)<br>362/385 | 47<br>(22, 74)<br>7/15 | 55<br>(50, 61)<br>171/309 | 5<br>(2, 10)<br>7/145 | 96<br>(92, 99)<br>171/179 | 80<br>(69, 89)<br>55/69<br>**p=0.008**[**] | 45<br>(40, 50)<br>174/390<br>**p=0.005**[**] | 20<br>(16, 26)<br>55/271<br>**p<0.001**[**] | 93<br>(88, 96)<br>174/188<br>p=0.230[**] |

| | | | | | | | | | | | | |
|---|---|---|---|---|---|---|---|---|---|---|---|---|
| CRP<br>≥5mg/l | 84<br>(75, 92)<br>74/88<br><br>p=0.122† | 49<br>(46, 53)<br>338/691<br><br>p=0.942† | 17<br>(14, 22)<br>74/427<br><br>p=0.380† | 96<br>(94, 98)<br>338/352<br><br>p=0.271† | 67<br>(39, 89)<br>10/15<br><br>p=0.269† | 60<br>55, 66)<br>176/294<br><br>p=0.261† | 8<br>(4, 14)<br>10/128<br><br>p=0.308† | 97<br>(94, 100)<br>176/181<br><br>p=0.385† | 88<br>(78, 95)<br>58/66<br>**p=0.043****<br>p=0.199† | 41<br>(36, 47)<br>147/360<br>**p<0.001****<br>p=0.296† | 21<br>(17, 27)<br>58/271<br>**p=0.001****<br>p=0.751† | 95<br>91, 98)<br>147/155<br>p=0.256**<br>p=0.390† |
| ≥10mg/l | 74<br>(64, 83)<br>65/88<br><br>p=0.895† | 64<br>(61, 68)<br>444/691<br><br>**p<0.001†** | 21<br>(17, 26)<br>65/312<br><br>**p=0.042†** | 95<br>(93, 97)<br>444/467<br><br>p=0.500† | 33<br>(12, 62)<br>5/15<br><br>p=0.456† | 72<br>(67, 77)<br>211/294<br><br>**p<0.001†** | 6<br>(2, 13)<br>5/88<br><br>p=0.775† | 95<br>(92, 98)<br>211/221<br><br>p=0.979† | 82<br>(71, 91)<br>54/66<br>**p<0.001****<br>p=0.756† | 59<br>(54, 65)<br>213/360<br>**p<0.01****<br>**p<0.001†** | 27<br>(21, 34)<br>54/201<br>**p<0.001****<br>p=0.094† | 95<br>(91, 98)<br>213/225<br>p=0.693**<br>p=0.379† |

Within row p-values: **CD4 count >350 vs. ≤350 cells/µl.
Within column p-values: *Any vs. ≥2-week cough, †W4SS vs. CRP.
Missing data or not done: no CD4 count within three months (n=76).
Abbreviations: CRP C-reactive protein; CI, confidence interval; NPV, negative predictive value; PPV, positive predictive value; TB, tuberculosis; W4SS, World Health Organization-recommended four-symptom screen.

**Supplementary Table 5**. Head-to-head diagnostic accuracy of haemoglobin (<10 g/dl) stratified by CD4 count and W4SS statuses. Haemoglobin had increased sensitivity at lower CD4 cells counts but decreased specificity (same trend observed in W4SS-positives vs. negatives). Non-head-to-head data are not shown due to the many patients not receiving haemoglobin testing (it was done programmatically). Data are %, 95% CI, and n/N.

| **All participants (n=634)** | | | | **CD4 count >350 cells/µl (n=243)** | | | | **CD4 count ≤350 cells/µl (n=360)** | | | |
|---|---|---|---|---|---|---|---|---|---|---|---|
| **Sensitivity** | **Specificity** | **PPV** | **NPV** | **Sensitivity** | **Specificity** | **PPV** | **NPV** | **Sensitivity** | **Specificity** | **PPV** | **NPV** |
| 31 | 88 | 27 | 90 | 8 | 91 | 5 | 95 | 37 | 86 | 35 | 86 |
| (22, 43) | (86, 91) | (19, 38) | (87, 93) | (1, 37) | (87, 95) | (1, 24) | (91, 98) | (25, 50) | (81, 90) | (24, 48) | (82, 91) |
| 25/81 | 487/553 | 25/91 | 487/543 | 1/13 | 210/230 | 1/21 | 210/222 | 23/63 | 254/297 | 23/66 | 254/294 |
| | | | | | | | | **p=0.042**[*] | **p=0.042**[*] | **p=0.007**[*] | **p=0.002**[*] |
| **p<0.001**[†] | **p<0.001**[†] | **p=0.041**[†] | p=0.062[†] | **p=0.013**[†] | **p<0.001** | p=0.784[†] | p=0.972[†] | **p<0.001**[†] | **p<0.001**[†] | p=0.055[†] | p=0.066[†] |
| **p<0.001**[‡] | **p<0.001**[‡] | p=0.519[‡] | **p=0.002**[‡] | **p=0.050**[‡] | **p<0.001**[‡] | p=0.630[‡] | p=0.830[‡] | **p<0.001**[‡] | **p<0.001**[‡] | p=0.435[‡] | **p=0.002**[‡] |
| | | | | **W4SS-positive (n=354)** | | | | **W4SS-negative (n=280)** | | | |
| | | | | 39 | 86 | 37 | 86 | 0 | 91 | 0 | 93 |
| | | | | (28, 53) | (81, 90) | (26, 50) | (82, 91) | (0, 20) | (87, 95) | (0, 15) | (90, 97) |
| | | | | 25/64 | 248/290 | 25/67 | 248/287 | 0/17 | 239/263 | 0/24 | 239/256 |
| | | | | | | | | **p=0.002**[*] | p=0.052[*] | **p<0.001**[*] | **p=0.008**[*] |
| | | | | **p<0.001**[‡‡] | **p<0.001**[‡‡] | p=0.255[‡‡] | **p<0.001**[‡‡] | **p=0.008**[‡‡] | **p<0.001**[‡‡] | p=0.132[‡‡] | p=0.761[‡‡] |

Within row p-values: [*]CD4 count >350 vs. ≤350 cells/µl.
Within column p-values: [†]haemoglobin vs. W4SS in **Table 2**, [‡]haemoglobin vs. CRP (≥10mg/l) in **Table 2** (comparisons within a CD4 count stratum); [‡‡] haemoglobin vs. CRP (≥10mg/l) in **Supplementary Table 1** (comparisons within a W4SS stratum).
Missing data or not done: CD4 count within three months (n=31); haemoglobin not programmatically done for 21% (166/800) of the people in the W4SS-CRP head-to-head analysis.
Abbreviations: CI, confidence interval; NPV, negative predictive value; PPV, positive predictive value; W4SS; World Health Organization-recommended four-symptom screen.

**Supplementary Table 6.** AUROCs, rule-in (~95% specificity), rule-out (~95% sensitivity), and Youden index (maximum sum of sensitivity and specificity) thresholds of W4SS, CRP, and haemoglobin, stratified by W4SS and CD4 count status. CRP had the highest AUROC. AUROCs were increased in W4SS-positives (vs. -negatives) and in people with CD4 counts >350 cells/µl (vs. ≤350). Haemoglobin followed similar trends. At rule-out thresholds, no tests met the WHO TPP minimal specificity point estimate of 70%, irrespective of patient group. Data are % (95% CI).

| | **W4SS** | | | **CRP** | | | **Haemoglobin** | | |
|---|---|---|---|---|---|---|---|---|---|
| | **Overall** | | | | | | | | |
| | AUROC: 0.70 (0.64, 0.75) | | | AUROC: 0.78 (0.73, 0.84) | | | AUROC: 0.70 (0.64, 0.75) | | |
| | Threshold (number) | Sensitivity | Specificity | Threshold (mg/l) | Sensitivity | Specificity | Threshold (g/dl) | Sensitivity | Specificity |
| Rule-in | ≥3 | 32 (25, 40) | 89 (87, 91) | >130 | 31 (20, 36) | 95 (93, 96) | <9 | 13 (8-21) | 95 (93, 97) |
| Rule-out | ≥0 | 100 (97, 100) | 0 (0, 0) | >3 | 89 (83, 94) | 39 (35, 42) | <14 | 95 (89, 98) | 20 (17, 23) |
| Youden's index | ≥2 | 62 (54, 70) | 73 (70, 76) | >18 | 75 (66, 82) | 75 (72, 77) | <13 | 82 (74, 89) | 51 (47, 54) |
| | | | | **W4SS-positives** | | | | | |
| | | | | AUROC: 0.82 (0.77, 0.87) | | | AUROC: 0.72 (0.65, 0.78) | | |
| Rule-in | | | | >172 | 30 (22, 40) | 95 (93, 97) | <9 | 23 (15, 33) | 95 (92, 97) |
| Rule-out | | | | >4 | 95 (89, 98) | 38 (34, 43) | <14 | 95 (89, 99) | 19 (15, 23) |
| Youden's index | | | | >18 | 89 (81, 94) | 65 (61, 69) | <12 | 71 (60, 80) | 65 (60, 69) |
| | | | | **W4SS-negatives** | | | | | |
| | | | | AUROC: 0.58 (0.45, 0.70) | | | AUROC: 0.56 (0.45, 0.68) | | |
| Rule-in | | | | >72 | 4 (0, 18) | 95 (92, 97) | <9 | 0 (0, 15) | 95 (92, 97) |
| Rule-out | | | | >3 | 67 (48, 82) | 46 (41, 51) | <14 | 94 (76, 100) | 27 (23, 32) |
| Youden's index | | | | >5 | 58 (40, 75) | 61 (56, 65) | Same as rule-out | | |
| | **CD4 count >350 cells/µl** | | | | | | | | |
| | AUROC: 0.58 (0.42, 0.73) | | | AUROC: 0.62 (0.47, 0.77) | | | AUROC: 0.59 (0.45, 0.74) | | |
| Rule-in | ≥3 | 93 (90, 95) | 22 (8, 44) | >91 | 95 (92, 97) | 17 (5, 38) | <10 | 0 (0, 21) | 95 (92, 97) |
| Rule-out | ≥0 | 100 (85, 100) | 0 (0, 1) | >6 | 67 (45, 84) | 64 (59, 68) | <14 | 92 (68, 100) | 23 (18, 78) |
| Youden's index | ≥2 | 22 (8, 44) | 93 (90, 95) | Same as rule-out | | | <12 | 54 (29, 78) | 68 (63, 73) |
| | **CD4 count ≤350 cells/µl** | | | | | | | | |
| | AUROC: 0.70 (0.64, 0.75) | | | AUROC: 0.81 (0.75, 0.86) | | | AUROC: 0.70 (0.63, 0.76) | | |
| Rule-in | ≥3 | 32 (25, 40) | 89 (87, 91) | >130 | 36 (26, 46) | 95 (92, 96) | <9 | 12 (6, 21) | 95 (93, 97) |
| Rule-out | ≥0 | 100 (97, 100) | 0 (0, 0) | >3 | 95 (88, 98) | 30 (26, 35) | <14 | 95 (89, 99) | 21 (18, 25) |
| Youden's index | ≥2 | 62 (54, 70) | 73 (70, 76) | >18 | 82 (73, 89) | 70 (65, 74) | <13 | 83 (74, 90) | 50 (45, 54) |

Abbreviations: AUROC, area under the receiver operator characteristic curve; CRP, C-reactive protein; CI, confidence interval; W4SS, WHO-recommended four-symptom screen; WHO, World Health Organization; WHO TTP, World Health Organization target product profile.

**Supplementary Table 7.** Head-to-head diagnostic accuracy of sputum Xpert and Ultra for TB stratified by CD4 cell count. Each test had higher sensitivities and similar specificities in people with lower vs. higher CD4 cell counts. Ultra had higher sensitivity than Xpert at CD4 counts ≤350 cells/µl. We did not sub-stratify CD4 cell counts categories further by smear status given low numbers of index test-detected cases. Data are %, 95% CI, and n/N.

| | **All (n=787)** | | | | **CD4 count >350 cells/µl (n=304)** | | | | **CD4 count ≤350 cells/µl (n=432)** | | | |
|---|---|---|---|---|---|---|---|---|---|---|---|---|
| | **Sensitivity** | **Specificity** | **PPV** | **NPV** | **Sensitivity** | **Specificity** | **PPV** | **NPV** | **Sensitivity** | **Specificity** | **PPV** | **NPV** |
| Smear microscopy | 15<br>(9, 24)<br>16/104 | 99<br>(98, 100)<br>678/683 | 76<br>(53, 92)<br>16/21 | 89<br>(86, 91)<br>678/766 | 18<br>(4, 44)<br>3/17 | 100<br>(99, 100)<br>287/287 | 100<br>(30, 100)<br>3/3 | 96<br>(93, 98)<br>287/301 | 13<br>(7, 23)<br>10/77<br>p=0.683[*] | 99<br>(97, 100)<br>350/355<br>**p=0.044[*]** | 67<br>(39, 89)<br>10/15<br>p=0.239[*] | 84<br>(81, 88)<br>350/417<br>**p<0.001[*]** |
| Xpert | 56<br>(46, 66)<br>58/104<br><br>**p<0.001[†]** | 99<br>(98, 100)<br>676/683<br><br>p=0.562[†] | 89<br>(79, 96)<br>58/65<br><br>p=0.134[†] | 94<br>(92, 95)<br>676/722<br><br>**p=0.001[†]** | 30<br>(11, 56)<br>5/17<br><br>p=0.419[†] | 99<br>(97, 100)<br>284/287<br><br>p=0.082[†] | 63<br>(25, 92)<br>5/8<br><br>p=0.214[†] | 96<br>(94, 98)<br>284/296<br><br>p=0.721[†] | 60<br>(48, 71)<br>46/77<br>**p=0.023[*]**<br>**p<0.001[†]** | 99<br>(98, 100)<br>351/355<br>p=0.921[*]<br>p=0.737[†] | 92<br>(81, 98)<br>46/50<br>**p=0.017[*]**<br>**p=0.013[†]** | 92<br>(89, 95)<br>351/382<br>**p=0.031[*]**<br>**p<0.001[†]** |
| Ultra-negative | 10<br>(2, 27)<br>3/30 | 99<br>(98, 100)<br>663/668 | 38<br>(9, 76)<br>3/8 | 96<br>(94, 97)<br>663/690 | 0<br>(0, 31)<br>0/10 | 99<br>97, 100)<br>280/283 | 0<br>0, 71)<br>0/3 | 97<br>(94, 98)<br>280/290 | 17<br>4, 41)<br>3/18<br>p=0.172[*] | 99<br>(98, 100)<br>342/344<br>p=0.502[*] | 60<br>(15, 95)<br>3/5<br>p=0.090[*] | 96<br>(93, 98)<br>342/357<br>p=0.621[*] |
| Ultra | 71<br>(61, 80)<br>74/104<br><br>**p=0.014[‡]** | 98<br>(96, 99)<br>668/683<br><br>p=0.086[‡] | 83<br>(74, 90)<br>74/89<br><br>p=0.287[‡] | 96<br>(94, 97)<br>668/698<br><br>p=0.083[‡] | 42<br>(19, 68)<br>7/17<br><br>p=0.473[‡] | 99<br>(97, 100)<br>283/287<br><br>p=0.704[‡] | 64<br>(31, 90)<br>7/11<br><br>p=0.960[‡] | 97<br>(94, 99)<br>283/293<br><br>p=0.682[‡] | 77<br>(66, 86)<br>59/77<br>**p=0.004[*]**<br>**p=0.025[‡]** | 97<br>(95, 99)<br>344/355<br>p=0.155[*]<br>p=0.068[‡] | 85<br>(74, 92)<br>59/70<br>p=0.101[*]<br>p=0.208[‡] | 96<br>(93, 98)<br>344/362<br>p=0.327[*]<br>p=0.084[‡] |
| Xpert-negative | 41<br>(27, 57)<br>19/46 | 98<br>(97, 99)<br>663/676 | 59<br>(41, 76)<br>19/32 | 96<br>(94, 97)<br>663/690 | 17<br>(3, 49)<br>2/12 | 99<br>(97, 100)<br>280/284 | 34<br>(5, 78)<br>2/6 | 97<br>(94, 99)<br>280/290 | 5<br>(34, 70)<br>16/31<br>**p=0.037[*]** | 98<br>(96, 99)<br>342/351<br>p=0.307[*] | 64<br>(43, 83)<br>16/25<br>p=0.172[*] | 96<br>(94, 98)<br>342/357<br>p=0.621[*] |

Within row p-values: [*]CD4 count ≤350 vs. >350 cells/µl.
Within column p-values: [†]smear vs. Xpert; [‡]Xpert vs. Ultra.
Missing data or not done: CD4 count (n=74).
Abbreviations: CI, confidence interval; NPV, negative predictive value; PPV, positive predictive value; TB, tuberculosis; Ultra, Xpert MTB/RIF Ultra; Xpert, Xpert MTB/RIF.

**Supplementary Table 8**. Diagnostic accuracy of Xpert and Ultra for TB stratified by W4SS status in smear-negative people. Trends were like those seen overall (**Table 4**). Data are %, 95% CI, and n/N.

| | **Head-to-head** | | | | | | | | | | | |
|---|---|---|---|---|---|---|---|---|---|---|---|---|
| | **All<br>(n=787)** | | | | **W4SS-positive<br>(n=357)** | | | | **W4SS-negative<br>(n=430)** | | | |
| | **Sensitivity** | **Specificity** | **PPV** | **Sensitivity** | **Specificity** | **PPV** | **Sensitivity** | **Specificity** | **PPV** | **Sensitivity** | **Specificity** | **PPV** |
| Xpert | 50<br>(39, 61)<br>44/88 | 99<br>(98, 100)<br>671/678 | 86<br>(74, 94)<br>44/51 | 94<br>(92, 95)<br>671/715 | 22<br>(8, 44)<br>5/23 | 100<br>(98, 100)<br>328/331 | 63<br>(25, 92)<br>5/8 | 95<br>(92, 97)<br>328/346 | 60<br>(48, 72)<br>39/65<br>**p=0.002**[*] | 99<br>(98, 100)<br>343/347<br>p=0.751[*] | 91<br>(78, 98)<br>39/43<br>**p=0.033**[*] | 93<br>(90, 96)<br>343/369<br>p=0.305[*] |
| Ultra | 67<br>(56, 77)<br>59/88<br>**p=0.022**[†] | 98<br>(96, 99)<br>663/678<br>p=0.089[†] | 80<br>(69, 88)<br>59/74<br>p=0.625[†] | 96<br>(94, 97)<br>663/692<br>p=0.117[†] | 44<br>(24, 66)<br>10/23<br>p=0.116[†] | 99<br>(97, 100)<br>327/331<br>p=0.704[†] | 72<br>(42, 92)<br>10/14<br>p=0.665[†] | 97<br>(94, 98)<br>327/340<br>p=0.385[†] | 76<br>(64, 86)<br>49/65<br>**p=0.005**[*]<br>p=0.061[†] | 97<br>(95, 99)<br>336/347<br>p=0.083[*]<br>p=0.068[†] | 82<br>(70, 91)<br>49/60<br>p=0.391[*]<br>p=0.200[†] | 96<br>(93, 98)<br>336/352<br>p=0.636<br>p=0.152[†] |
| **Non-head-to-head** | | | | | | | | | | | | |
| Xpert | 50<br>(39, 61)<br>44/88 | 99<br>(98, 100)<br>672/679 | 86<br>(74, 94)<br>44/51 | 94<br>(92, 95)<br>672/716 | 22<br>(7, 44)<br>5/23 | 99<br>(97, 100)<br>328/331 | 63<br>(24, 91)<br>5/8 | 95<br>(92, 97)<br>328/346 | 60<br>(47, 72)<br>39/65<br>**p=0.002**[*] | 99<br>(97, 100)<br>344/348<br>p=0.754[*] | 91<br>(78, 97)<br>39/43<br>**p=0.033**[*] | 93<br>(90, 95)<br>344/370<br>p=0.310[*] |
| Ultra | 68<br>(58, 78)<br>62/91<br>**p=0.014**[†] | 98<br>(97, 99)<br>725/741<br>p=0.092[†] | 79<br>(69, 88)<br>62/78<br>p=0.325[†] | 96<br>(95, 97)<br>725/754<br>**p=0.043**[†] | 76<br>(65, 86)<br>52/68<br>**p<0.001**[†] | 97<br>(95, 98)<br>367/379<br>**p=0.037**[†] | 81<br>(70, 90)<br>52/64<br>p=0.218[†] | 96<br>(93, 98)<br>367/383<br>p=0.512 [†] | 43<br>(23, 66)<br>10/23<br>**p=0.003**[*]<br>p=0.170[†] | 99<br>(97, 100)<br>358/362<br>p=0.794[*]<br>p=0.995[†] | 71<br>(42, 92)<br>10/14<br>**p<0.001**[*]<br>p=0.071[†] | 96<br>(94, 98)<br>358/371<br>p=0.631[*]<br>**p=0.032**[†] |

Within row p-values: [*]W4SS-positive vs. -negative.
Within column p-values: [†]Xpert vs. Ultra.
Abbreviations: CI, confidence interval; NPV, negative predictive value; PPV, positive predictive value; Ultra, W4SS, WHO four-symptom screen; Xpert MTB/RIF; Xpert, Xpert MTB/RIF Ultra.

**Supplementary Table 9**. Non-head-to-head diagnostic accuracy of Xpert and Ultra for TB detection stratified by CD4 count. Non-head-to-head diagnostic accuracy trends were like that in head-to-head comparisons (**Supplementary Table 7**). Data are %, 95% CI, and n/N.

| | **All participants** | | | | **CD4 count >350 cells/µl** | | | | **CD4 count ≤350 cells/µl** | | | |
|---|---|---|---|---|---|---|---|---|---|---|---|---|
| | **Sensitivity** | **Specificity** | **PPV** | **NPV** | **Sensitivity** | **Specificity** | **PPV** | **NPV** | **Sensitivity** | **Specificity** | **PPV** | **NPV** |
| Smear microscopy | 15<br>(9, 24)<br>16/107 | 100<br>(99, 100)<br>743/748 | 77<br>(53, 92)<br>16/21 | 90<br>(87, 92)<br>743/834 | 17<br>(4, 42)<br>3/18 | 100<br>(99, 100)<br>309/309 | 10<br>(30, 100)<br>3/3 | 95<br>(93, 98)<br>309/324 | 13<br>(7, 23)<br>10/79<br>p=0.406[*] | 99<br>(98, 100)<br>390/395<br>**p=0.044**[*] | 67<br>(39, 89)<br>10/15<br>p=0.197[*] | 85<br>(82, 89)<br>390/459<br>**p<0.001**[*] |
| Xpert | 56<br>(46, 66)<br>58/104<br>**p<0.001**[†] | 99<br>(98, 100)<br>677/684<br>p=0.462[†] | 90<br>(80, 96)<br>58/65<br>p=0.134[†] | 94<br>(92, 96)<br>677/723<br>**p=0.002**[†] | 29<br>(11, 56)<br>5/17<br>p=0.369[†] | 99<br>(97, 100)<br>284/287<br>p=0.072[†] | 63<br>(25, 92)<br>5/8<br>p=0.214[†] | 96<br>(94, 98)<br>284/296<br>p=0.726[†] | 60<br>(48, 71)<br>46/77<br>**p=0.023**[*]<br>**p<0.001**[†] | 99<br>(98, 100)<br>352/356<br>p=0.924[*]<br>p=0.858[†] | 92<br>(81, 98)<br>46/50<br>**p=0.017**[*]<br>**p=0.013**[†] | 92<br>(89, 95)<br>352/383<br>**p=0.032**[*]<br>**p=0.002**[†] |
| Smear-negative | 50<br>(40, 61)<br>44/88 | 99<br>(98, 100)<br>672/679 | 87<br>(74, 95)<br>44/51 | 94<br>(92, 96)<br>672/716 | 21<br>(5, 51)<br>3/14 | 99<br>(97, 100)<br>284/287 | 50<br>(12, 89)<br>3/6 | 96<br>(94, 99)<br>284/295 | 55<br>(43, 68)<br>37/67<br>**p=0.021**[*] | 99<br>(98, 100)<br>347/351<br>p=0.909[*] | 90<br>(77, 98)<br>37/41<br>**p=0.010**[*] | 92<br>(89, 95)<br>347/377<br>**p=0.023**[*] |
| Ultra-negative | 10<br>(3, 27)<br>3/30 | 100<br>(99, 100)<br>663/668 | 38<br>(9, 76)<br>3/8 | 97<br>(95, 98)<br>663/690 | 0<br>(0, 31)<br>0/10 | 99<br>(97, 100)<br>280/283 | 0<br>(0, 71)<br>0/3 | 97<br>(94, 99)<br>280/290 | 17<br>(4, 42)<br>3/18<br>p=0.172[*] | 99<br>(98, 100)<br>342/344<br>p=0.502[*] | 60<br>(15, 95)<br>3/5<br>p=0.090[*] | 96<br>(94, 98)<br>342/357<br>p=0.621[*] |
| Ultra | 72<br>(63, 81)<br>77/107<br>**p=0.017**[‡] | 97<br>(95, 98)<br>730/760<br>**p<0.001**[‡] | 83<br>(74, 90)<br>77/93<br>p=0.259[‡] | 97<br>(95, 98)<br>730/760<br>**p=0.04**[‡] | 44<br>(22, 70)<br>8/18<br>p=0.358[‡] | 99<br>(97, 100)<br>305/309<br>p=0.778[‡] | 67<br>(35, 91)<br>8/12<br>p=0.848[‡] | 97<br>(95, 99)<br>305/315<br>p=0.560[‡] | 77<br>(67, 86)<br>61/79<br>**p=0.006**[*]<br>**p=0.019**[‡] | 97<br>(95, 99)<br>381/393<br>p=0.121[*]<br>p=0.068[‡] | 84<br>(74, 92)<br>61/73<br>p=0.165[*]<br>p=0.172[‡] | 95<br>(93, 98)<br>381/39<br>p=0.361[*]<br>**p=0.039**[‡] |
| Smear-negative | 69<br>(58, 78)<br>62/91<br>**p=0.010**[‡] | 98<br>(97, 99)<br>725/741<br>p=0.092[‡] | 80<br>(69, 88)<br>62/78<br>p=0.325[‡] | 97<br>(95, 98)<br>725/754<br>**p=0.035**[‡] | 40<br>(17, 68)<br>6/15<br>p=0.208[‡] | 99<br>(97, 100)<br>305/309<br>p=0.778[‡] | 60<br>(27, 88)<br>6/10<br>p=0.696[‡] | 97<br>(95, 99)<br>305/314<br>p=0.551[‡] | 74<br>(62, 84)<br>51/69<br>**p=0.011**[*]<br>**p=0.023**[‡] | 97<br>(95, 99)<br>376/388<br>p=0.115[*]<br>p=0.068[‡] | 81<br>(70, 90)<br>51/63<br>p=0.137[*]<br>p=0.199[‡] | 95<br>(93, 98)<br>376/394<br>p=0.240[*]<br>p=0.052[‡] |
| Xpert-negative | 42<br>(27, 57)<br>19/46<br>**p=0.003**[‡] | 99<br>(97, 99)<br>663/676<br>p=0.061[‡] | 60<br>(41, 77)<br>19/32<br>p=0.266[‡] | 97<br>(95, 98)<br>663/690<br>p>0.999[‡] | 17<br>(3, 49)<br>2/12<br>p=0.176[‡] | 99<br>(97, 100)<br>280/284<br>p=0.707[‡] | 33<br>(5, 78)<br>2/6<br>p=0.257[‡] | 97<br>(94, 99)<br>280/290<br>p>0.999[‡] | 52<br>(34, 70)<br>16/31<br>**p=0.037**[*]<br>**p=0.016**[‡] | 97<br>(96, 99)<br>342/351<br>p=0.307[*]<br>p=0.036[‡] | 64<br>(43, 83)<br>16/25<br>p=0.172[*]<br>p=0.865[‡] | 96<br>(94, 98)<br>342/357<br>p=0.621[*]<br>p>0.999[‡] |

Within row p-values: [*] CD4 counts ≤350 vs. >350 cells/µl.
Within column p-values: [†]smear vs. Xpert and [‡]Xpert vs. Ultra.
Abbreviations: CI, confidence interval; NPV, negative predictive value; PPV, positive predictive value; Ultra, Xpert MTB/RIF Ultra; Xpert, Xpert MTB/RIF.

**Supplementary Table 10**. Non-head-to-head diagnostic accuracy of Xpert and Ultra for TB overall, and stratified by W4SS status. These findings are like those in the head-to-head analysis (**Table 4**). Data are %, 95% CI, and n/N.

| | All | | | | W4SS-negative | | | | W4SS-positive | | | |
|---|---|---|---|---|---|---|---|---|---|---|---|---|
| | **Sensitivity** | **Specificity** | **PPV** | **NPV** | **Sensitivity** | **Specificity** | **PPV** | **NPV** | **Sensitivity** | **Specificity** | **PPV** | **NPV** |
| Smear microscopy | 15<br>(9, 24)<br>16/107 | 100<br>(99, 100)<br>743/748 | 77<br>(53, 92)<br>16/21 | 90<br>(87, 92)<br>743/834 | 12<br>(3, 31)<br>3/26 | 100<br>(99, 100)<br>362/362 | 100<br>(30, 100)<br>3/3 | 95<br>(92, 97)<br>362/385 | 17<br>(9, 26)<br>13/81<br>p=0.575$^*$ | 99<br>(98, 100)<br>381/386<br>**p=0.029**$^*$ | 73<br>(47, 91)<br>13/18<br>p=0.296$^*$ | 85<br>(82, 89)<br>381/449<br>**p<0.001**$^*$ |
| Xpert | 56<br>(46, 66)<br>58/104<br>**p<0.001**$^\dagger$ | 99<br>(98, 100)<br>677/684<br>p=0.462$^\dagger$ | 90<br>(80, 96)<br>58/65<br>p=0.134$^\dagger$ | 94<br>(92, 96)<br>677/723<br>**p<0.01**$^\dagger$ | 27<br>(12, 48)<br>7/26<br>p=0.159$^\dagger$ | 100<br>(98, 100)<br>328/331<br>p=0.069$^\dagger$ | 70<br>(35, 94)<br>7/10<br>p=0.279$^\dagger$ | 95<br>(92, 97)<br>328/347<br>p=0.772$^\dagger$ | 66<br>(54, 76)<br>51/78<br>**p=0.001**$^*$<br>**p<0.001**$^\dagger$ | 99<br>(98, 100)<br>349/353<br>p=0.768$^*$<br>p=0.841$^\dagger$ | 93<br>(83, 98)<br>51/55<br>**p=0.033**$^*$<br>**p=0.022**$^\dagger$ | 93<br>(90, 96)<br>349/376<br>p=0.348$^*$<br>**p<0.001**$^\dagger$ |
| Ultra-negative | 10<br>(3, 27)<br>3/30 | 100<br>(99, 100)<br>663/668 | 38<br>(9, 76)<br>3/8 | 97<br>(95, 98)<br>663/690 | 0<br>(0, 24)<br>0/14 | 100<br>(98, 100)<br>324/327 | 0<br>(0, 71)<br>0/3 | 96<br>(94, 98)<br>324/338 | 19<br>(5, 46)<br>3/16<br>p=0.088$^*$ | 100<br>(98, 100)<br>339/341<br>p=0.620$^*$ | 60<br>(15, 95)<br>3/5<br>p=0.090$^*$ | 97<br>(94, 99)<br>339/352<br>p=0.761$^*$ |
| Ultra | 72<br>(63, 81)<br>77/107<br>**p=0.014**$^\ddagger$ | 97<br>(95, 98)<br>730/760<br>**p<0.001**$^\ddagger$ | 83<br>(74, 90)<br>77/93<br>p=0.259$^\ddagger$ | 97<br>(95, 98)<br>730/760<br>**p=0.035**$^\ddagger$ | 47<br>(27, 67)<br>12/26<br>p=0.150$^\ddagger$ | 99<br>(98, 100)<br>358/362<br>p=0.794$^\ddagger$ | 75<br>(48, 93)<br>12/16<br>p=0.780$^\ddagger$ | 97<br>(94, 98)<br>358/372<br>p=0.273$^\ddagger$ | 81<br>(70, 89)<br>65/81<br>**p=0.001**$^*$<br>**p=0.035**$^\ddagger$ | 97<br>(95, 99)<br>372/384<br>p=0.057$^*$<br>p=0.064$^\ddagger$ | 85<br>(75, 92)<br>65/77<br>p=0.364$^*$<br>p=0.149$^\ddagger$ | 96<br>(94, 98)<br>372/388<br>p=0.799$^*$<br>p=0.607$^\ddagger$ |
| Xpert-negative | 42<br>(27, 57)<br>19/46<br>**p=0.003**$^\ddagger$ | 99<br>(97, 99)<br>663/676<br>p=0.061$^\ddagger$ | 60<br>(41, 77)<br>19/32<br>p=0.266$^\ddagger$ | 97<br>(95, 98)<br>663/690<br>p>0.999$^\ddagger$ | 27<br>(10, 52)<br>5/19<br>**p=0.037**$^\ddagger$ | 99<br>(97, 100)<br>324/328<br>p=0.707$^\ddagger$ | 56<br>(22, 87)<br>5/9<br>p=0.091$^\ddagger$ | 96<br>(94, 98)<br>324/338<br>p>0.999$^\ddagger$ | 52<br>(32, 72)<br>14/27<br>p=0.083$^*$<br>**p=0.032**$^\ddagger$ | 98<br>(96, 99)<br>339/348<br>p=0.196$^*$<br>**p=0.036**$^\ddagger$ | 61<br>(39, 81)<br>14/23<br>p=0.783$^*$<br>p=0.971$^\ddagger$ | 97<br>(94, 99)<br>339/352<br>p=0.761$^*$<br>p>0.999$^\ddagger$ |

Within row p-values: $^*$W4SS-positive vs. -negative.
Within column p-values: $^\dagger$smear vs. Xpert or $^\ddagger$Xpert vs. Ultra.
Abbreviations: CI, confidence interval; NPV, negative predictive value; PPV, positive predictive value; TB, tuberculosis; Ultra, Xpert MTB/RIF Ultra; W4SS, WHO four-symptom screen; WHO, World Health Organization; Xpert, Xpert MTB/RIF.

**Supplementary Table 11**. Head-to-head diagnostic accuracy of Xpert and Ultra for TB detection by previous TB status. No specificity differences occurred when data stratified by previous TB status. Data are %, 95% CI, and n/N.

| | **All (n=787)** | | | | **No previous TB (n=675)** | | | | **Previous TB (n=112)** | | | |
|---|---|---|---|---|---|---|---|---|---|---|---|---|
| | **Sensitivity** | **Specificity** | **PPV** | **NPV** | **Sensitivity** | **Specificity** | **PPV** | **NPV** | **Sensitivity** | **Specificity** | **PPV** | **NPV** |
| Xpert | 56<br>(46, 66)<br>58/104 | 99<br>(98, 100)<br>676/683 | 89<br>(79, 96)<br>58/65 | 94<br>(92, 95)<br>676/722 | 58<br>(47, 69)<br>51/88 | 99<br>(98, 100)<br>581/587 | 90<br>(79, 97)<br>51/57 | 95<br>(92, 96)<br>581/618 | 44<br>(20, 70)<br>7/16<br>p=0.293* | 99<br>(94, 100)<br>95/96<br>p=0.986* | 88<br>(47, 100)<br>7/8<br>p=0.866* | 91<br>(84, 96)<br>95/104<br>p=0.303* |
| Smear-negative | 50<br>(39, 61)<br>44/88 | 99<br>(98, 100)<br>671/678 | 86<br>(74, 94)<br>44/51 | 94<br>(92, 95)<br>671/715 | 54<br>(42, 65)<br>40/75 | 99<br>(98, 100)<br>576/582 | 87<br>(74, 96)<br>40/46 | 95<br>(93, 96)<br>576/611 | 31<br>(9, 61)<br>4/13<br>p=0.133* | 99<br>(94, 100)<br>95/96<br>p=0.992* | 80<br>(28, 99)<br>4/5<br>p=0.668* | 91<br>(84, 96)<br>95/104<br>p=0.251* |
| Ultra-negative | 10<br>(2, 27)<br>3/30 | 99<br>(98, 100)<br>663/668 | 38<br>(9, 76)<br>3/8 | 96<br>(94, 97)<br>663/690 | 13<br>(3, 33)<br>3/24 | 100<br>(98, 100)<br>570/575 | 38<br>(9, 76)<br>3/8 | 97<br>(95, 98)<br>570/591 | 0<br>(0, 46)<br>0/6<br>p=0.361* | 100<br>(96, 100)<br>93/93<br>p=0.367* | Non-calculable<br>- | 94<br>(87, 98)<br>93/99<br>p=0.234* |
| Ultra | 71<br>(61, 80)<br>74/104<br>**p=0.021‡** | 98<br>(96, 99)<br>668/683<br>p=0.086‡ | 83<br>(74, 90)<br>74/89<br>p=0.287‡ | 96<br>(94, 97)<br>668/698<br>p=0.083‡ | 73<br>(63, 82)<br>64/88<br>**p=0.039‡** | 98<br>(97, 99)<br>575/587<br>p=0.154‡ | 85<br>(75, 92)<br>64/76<br>p=0.380‡ | 96<br>(95, 98)<br>575/599<br>p=0.113‡ | 63<br>(35, 85)<br>10/16<br>p=0.406*<br>p=0.288‡ | 97<br>(91, 99)<br>93/96<br>p=0.503*<br>p=0.312‡ | 77<br>(46, 95)<br>10/13<br>p=0.517*<br>p=0.549‡ | 94<br>(87, 98)<br>93/99<br>p=0.351*<br>p=0.480‡ |
| Smear-negative | 67<br>(56, 77)<br>59/88<br>**p=0.022‡** | 98<br>(96, 99)<br>668/683<br>p=0.086‡ | 83<br>(74, 90)<br>74/89<br>p=0.625‡ | 96<br>(94, 97)<br>668/698<br>p=0.718‡ | 70<br>(58, 80)<br>52/75<br>**p=0.044‡** | 98<br>(97, 99)<br>570/582<br>p=0.154‡ | 82<br>(70, 90)<br>52/64<br>p=0.425‡ | 97<br>(95, 98)<br>570/593<br>p=0.134‡ | 54<br>(25, 81)<br>7/13<br>p=0.273*<br>p=0.234‡ | 97<br>(91, 99)<br>93/96<br>p=0.512*<br>p=0.312‡ | 70<br>(35, 93)<br>7/10<br>p=0.411*<br>p=0.680‡ | 94<br>(87, 98)<br>93/99<br>p=0.316*<br>p=0.480‡ |
| Xpert-negative | 41<br>(27, 57)<br>19/46<br>**p=0.003‡** | 98<br>(97, 99)<br>663/676<br>p=0.061‡ | 59<br>(41, 76)<br>19/32<br>p=0.266‡ | 96<br>(94, 97)<br>663/690<br>p>0.999‡ | 44<br>(28, 61)<br>16/37<br>**p=0.011‡** | 99<br>(97, 100)<br>570/581<br>p=0.136‡ | 60<br>(39, 78)<br>16/27<br>p=0.287‡ | 97<br>(95, 98)<br>570/591<br>p>0.999‡ | 33<br>(7, 70)<br>3/9<br>p=0.588*<br>p=0.114‡ | 98<br>(93, 100)<br>93/95<br>p=0.889*<br>p=0.160‡ | 60<br>(15, 95)<br>3/5<br>p=0.975*<br>- | 94<br>(87, 98)<br>93/99<br>p=0.234*<br>p>0.999‡ |

Within row p-values: *no previous TB vs. previous TB.
Within column p-values: ‡Xpert vs. Ultra.
Abbreviations: CI, confidence interval; NPV, negative predictive value; PPV, positive predictive value; TB, tuberculosis; Ultra, Xpert MTB/RIF; Xpert, Xpert MTB/RIF.

**Supplementary Table 12.** Effect of different trace re-categorisation strategies (reclassification or exclusion) on Ultra diagnostic accuracy, with stratification by previous TB status. Both the re-calculated estimate and change (Δ) are shown. In people without previous TB, re-categorising traces decreased sensitivity (reclassification only) and increases specificity (reclassification and exclusion strategies), while no differences were seen in people with previous TB. Data are %, 95% CI, and n/N.

| | **All (n=787)** | | | **No previous TB (n= 675)** | | | **Previous TB (n=112)** | | |
|---|---|---|---|---|---|---|---|---|---|
| | **Overall** | **Trace reclassified** | **Trace excluded** | **Overall** | **Trace reclassified** | **Trace excluded** | **Overall** | **Trace reclassified** | **Trace excluded** |
| Sensitivity | 71<br>(61, 80)<br>74/104 | 66<br>(57, 76)<br>69/104<br>-5 (-10, 0)<br>p=0.454[†] | 70<br>(60, 79)<br>69/99<br>-1 (-11, 14)<br>p=0.820[†]<br>p=0.609[‡] | 73<br>(63, 82)<br>64/88 | 67<br>(57, 77)<br>59/88<br>-6 (-12, 0)<br>p=0.411[†] | 71<br>(61, 81)<br>59/83<br>-2 (-15, 12)<br>p=0.811[†]<br>p=0.568[‡] | 63<br>(35, 85)<br>10/16<br>p=0.406[*] | 63<br>(35, 85)<br>10/16<br>0 (-6, 6)<br>p=0.723[*]<br>p>0.999[†] | 63<br>(35, 85)<br>10/16<br>0 (-6, 6)<br>p=0.494[*]<br>p>0.999[†]<br>p>0.999[‡] |
| Specificity | 98<br>(96, 99)<br>668/683 | 99<br>(99, 100)<br>679/683<br>1 (1, 3)<br>**p=0.011[†]** | 99<br>(99, 100)<br>668/672<br>1 (0, 3)<br>**p=0.012[†]**<br>p=0.982[‡] | 98<br>(97, 99)<br>575/587 | 99<br>(99, 100)<br>584/587<br>2 (0, 3)<br>**p=0.019[†]** | 99<br>(99, 100)<br>575/578<br>2 (0, 3)<br>**p=0.021[†]**<br>p=0.985[‡] | 97<br>(91, 99)<br>93/96<br>p=0.503[*] | 99<br>(94, 100)<br>95/96<br>2 (-2, 6)<br>p=0.528[*]<br>p=0.157[†] | 99<br>(94, 100)<br>93/94<br>(2, -2, 6)<br>p=0.524[*]<br>p=0.322[†]<br>p=0.988[‡] |
| PPV | 83<br>(74, 90)<br>74/89 | 95<br>(87, 99)<br>69/73<br>11 (2, 21)<br>**p=0.025[†]** | 95<br>(87, 99)<br>69/73<br>11 (2, 21)<br>**p=0.025[†]**<br>p>0.999[‡] | 85<br>(75, 92)<br>64/76 | 95<br>(87, 99)<br>59/62<br>11 (1, 21)<br>**p=0.040[†]** | 95<br>(87, 99)<br>59/62<br>11 (1, 21)<br>**p=0.004[†]**<br>p>0.999[‡] | 77<br>(46, 95)<br>10/13<br>p=0.517[*] | 91<br>(59, 100)<br>10/11<br>13 (-15, 43)<br>p=0.568[*]<br>p=0.360[†] | 91<br>(59, 100)<br>10/11<br>13 (-15, 43)<br>p=0.568[*]<br>p=0.360[†]<br>p>0.999[‡] |
| NPV | 96<br>(94, 97)<br>668/698 | 95<br>(94, 97)<br>679/714<br>-1 (-3, 2)<br>p=0.588[†] | 96<br>(94, 98)<br>668/698<br>0 (-2, 2)<br>p>0.999[†]<br>p=0.588[‡] | 96<br>(95, 98)<br>575/599 | 95<br>(94, 97)<br>584/613<br>-1 (-3, 2)<br>p=0.538[†] | 96<br>(95, 98)<br>575/599<br>p>0.999[†]<br>p=0.538[‡] | 94<br>(87, 98)<br>93/99<br>p=0.351[*] | 94<br>(88, 98)<br>95/101<br>0 (-6, 6)<br>p=0.602[*]<br>p=0.971[†] | 94<br>(87, 98)<br>93/99<br>0 (-7, 7)<br>p=0.351[*]<br>p>0.999[†]<br>p=0.971[‡] |

Within row p-values: [*]No previous TB vs. previous TB; [†]vs. overall in participants of the same previous TB status; [‡]trace reclassified vs. excluded.
Abbreviations: CI, confidence interval; NPV, negative predictive value; PPV, positive predictive value; TB, tuberculosis; Ultra, Xpert MTB/RIF Ultra.

**Supplementary Table 13.** Diagnostic accuracy of urine tests (LF-LAM, concentrated Ultra) for TB overall and by W4SS status (head-to-head and non-head-to-head data). Urine Ultra and LF-LAM had similar sensitivity and specificity overall and by W4SS. A positive Ultra had higher PPV in W4SS-positives vs. -negatives, including in LF-LAM-negatives. Similarly, Ultra and LF-LAM had higher NPVs in W4SS-negatives vs. positives. Data are %, 95% CI, and n/N.

| | **Head-to-head** | | | | | | | | | | | |
|---|---|---|---|---|---|---|---|---|---|---|---|---|
| | **All participants (n=732)** | | | | **W4SS-negative (n=339)** | | | | **W4SS-positive (n=393)** | | | |
| | **Sensitivity** | **Specificity** | **PPV** | **NPV** | **Sensitivity** | **Specificity** | **PPV** | **NPV** | **Sensitivity** | **Specificity** | **PPV** | **NPV** |
| LF-LAM | 15<br>(9, 24)<br>14/97 | 99<br>(98, 100)<br>623/635 | 61<br>(39, 81)<br>14/23 | 89<br>(86, 91)<br>626/709 | 9<br>(2, 29)<br>2/23 | 100<br>(98, 100)<br>313/316 | 40<br>(6, 86)<br>2/5 | 94<br>(91, 97)<br>313/334 | 17<br>(9, 27)<br>12/74<br>p=0.370$^{*}$ | 99<br>(96, 100)<br>313/319<br>p=0.321$^{*}$ | 67<br>(41, 87)<br>12/18<br>p=0.280$^{*}$ | 84<br>(80, 88)<br>313/375<br>**p<0.001**$^{*}$ |
| Conc. Ultra-negatives | 9<br>(4, 18)<br>6/73 | 99<br>(98, 100)<br>619/628 | 40<br>(17, 68)<br>6/15 | 91<br>(88, 93)<br>619/686 | 5<br>(1,25)<br>1/20 | 100<br>(98, 100)<br>309/312 | 25<br>(1, 81)<br>1/4 | 95<br>(92, 97)<br>309/328 | 10<br>(4,21)<br>5/53<br>p=0.538$^{*}$ | 9<br>(96, 100)<br>310/316<br>p=0.323$^{*}$ | 46<br>(17, 77)<br>5/11<br>p=0.475$^{*}$ | 87<br>(83,90)<br>310/358<br>**p<0.001**$^{*}$ |
| Conc. Ultra | 25<br>(17, 35)<br>24/97<br>p=0.070$^{**}$ | 99<br>(98, 100)<br>628/635<br>p=0.248$^{**}$ | 78<br>(59, 91)<br>24/31<br>p=0.188$^{**}$ | 90<br>(88, 92)<br>628/701<br>p=0.439$^{**}$ | 14<br>(3, 34)<br>3/23<br>p=0.636$^{**}$ | 99<br>(97, 100)<br>312/316<br>p=0.704$^{**}$ | 43<br>(10, 82)<br>3/7<br>p=0.921$^{**}$ | 94<br>(91, 97)<br>312/332<br>p=0.888$^{**}$ | 29<br>(19,41)<br>21/74<br>p=0.137$^{*}$<br>p=0.076$^{**}$ | 100<br>(98, 100)<br>316/319<br>p=0.695$^{*}$<br>p=0.314$^{**}$ | 88<br>(68, 98)<br>21/24<br>**p=0.013**$^{*}$<br>p=0.103$^{**}$ | 86<br>(82, 90)<br>316/369<br>**p<0.001**$^{*}$<br>p=0.413$^{**}$ |
| LF-LAM-negatives | 20<br>(12, 30)<br>16/83<br>**p=0.048**$^{\dagger}$ | 99<br>(98, 100)<br>619/626<br>p=0.619$^{\dagger}$ | 70<br>(48, 87)<br>16/23<br>p=0.071$^{\dagger}$ | 91<br>(88, 93)<br>619/686<br>p>0.999$^{\dagger}$ | 10<br>(2, 31)<br>2/21<br>p=0.578$^{\dagger}$ | 99<br>(97, 100)<br>309/313<br>p=0.707$^{\dagger}$ | 34<br>(5, 78)<br>2/6<br>p=0.778$^{\dagger}$ | 95<br>(92, 97)<br>309/328<br>p>0.999$^{\dagger}$ | 23<br>(13, 35)<br>14/62<br>p=0.190$^{*}$<br>p=0.058$^{\dagger}$ | 100<br>(98, 100)<br>310/313<br>p=0.694$^{*}$<br>p=0.321$^{\dagger}$ | 83<br>(57, 97)<br>14/17<br>**p=0.025**$^{*}$<br>**p=0.041**$^{\dagger}$ | 87<br>(83, 90)<br>310/358<br>**p<0.001**$^{*}$<br>p>0.999$^{\dagger}$ |
| | **Non-head-to-head** | | | | | | | | | | | |
| LF-LAM | 16<br>(9, 4)<br>16/106 | 99<br>(98, 100)<br>732/745 | 56<br>(36, 74)<br>16/29 | 90<br>(87, 92)<br>732/822 | 8<br>(1, 26)<br>2/26 | 99<br>(97, 100)<br>356/362 | 25<br>(4, 66)<br>2/8 | 94<br>(91, 96)<br>356/380 | 18<br>(10, 28)<br>14/80<br>p=0.225$^{*}$ | 99<br>(97, 100)<br>376/383<br>p=0.859$^{*}$ | 67<br>(44, 86)<br>14/21<br>**p=0.044**$^{*}$ | 86<br>(82, 89)<br>376/442<br>**p<0.001**$^{*}$ |
| Conc. Ultra-negatives | 9<br>(4, 18)<br>6/73 | 99<br>(98, 100)<br>732/745 | 34<br>(14, 60)<br>6/18 | 91<br>(88, 93)<br>616/683 | 5<br>(1, 25)<br>1/20 | 99<br>(97, 100)<br>356/362 | 15<br>(1, 58)<br>1/7 | 95<br>(92, 97)<br>306/325 | 10<br>(4, 21)<br>5/53<br>p=0.538$^{*}$ | 99<br>(97, 100)<br>376/383<br>p=0.859$^{*}$ | 46<br>(17, 77)<br>5/11<br>p=0.171$^{*}$ | 87<br>(83, 90)<br>310/358<br>**p=0.001**$^{*}$ |
| Conc. Ultra | 25<br>(17, 35)<br>24/97<br>p=0.084$^{**}$ | 99<br>(98, 100)<br>629/636<br>p=0.318$^{**}$ | 78<br>(59, 91)<br>24/31<br>p=0.068$^{**}$ | 90<br>(88, 92)<br>629/702<br>p=0.729$^{**}$ | 14<br>(3, 34)<br>3/23<br>p=0.537$^{**}$ | 99<br>(97, 100)<br>312/316<br>p=0.673$^{**}$ | 43<br>(10, 382)<br>3/7<br>p=0.464$^{**}$ | 94<br>(91,9 7)<br>312/332<br>p=0.872$^{**}$ | 29<br>(19, 41)<br>21/74<br>p=0.137$^{*}$<br>p=0.108$^{**}$ | 100<br>(98, 100)<br>317/320<br>p=0.692$^{*}$<br>p=0.321$^{**}$ | 88<br>(68, 98)<br>21/24<br>**p=0.013**$^{*}$<br>p=0.094$^{**}$ | 86<br>(82, 90)<br>317/370<br>**p<0.001**$^{*}$<br>p=0.807$^{**}$ |
| LF-LAM-negatives | 20<br>(12, 30)<br>16/83 | 99<br>(98, 100)<br>616/623 | 70<br>(48, 87)<br>16/23 | 91<br>(88, 93)<br>616/683 | 10<br>(2, 31)<br>2/21 | 99<br>(97, 100)<br>306/310 | 34<br>(5,78)<br>2/6 | 95<br>(92, 97)<br>306/325 | 23<br>(13, 35)<br>14/62<br>p=0.190$^{*}$ | 100<br>(98, 100)<br>310/313<br>p=0.694$^{*}$ | 83<br>(57, 97)<br>14/17<br>**p=0.025**$^{*}$ | 87<br>(83, 90)<br>310/358<br>**p<0.001**$^{*}$ |

| | | | | | | | | | | | | |
|---|---|---|---|---|---|---|---|---|---|---|---|---|
| | **p=0.048†** | p=0.340† | **p=0.021†** | p>0.999† | p=0.578† | p=0.695† | p=0.416† | p>0.999 † | p=0.058† | p=0.338† | **p=0.041†** | p>0.999† |

Within row p-values: *W4SS-positive vs. -negatives.
Within column p-values: **LF-LAM vs. conc. Ultra; †LF-LAM in Ultra-negatives vs. Ultra in LF-LAM-negatives.
Abbreviations: CI, confidence interval; Conc., concentrated; LF-LAM, Determine TB LAM Ag test (LF-LAM), NPV, negative predictive value; PPV, positive predictive value; TB, tuberculosis, Ultra, Xpert MTB/RIF Ultra; W4SS, WHO-recommended four-symptom screen.

**Supplementary Table 14.** Number of people who could expectorate different numbers of sputum specimens (≥1 ml each). Whether people could naturally expectorate sputum before induction was offered was only successfully recorded for the last 163 patients (**Methods**) and, of these, five did not have any culture attempted due to human error and are omitted (the number of sputa induction successfully produced are specified in the footnote). The culture-positivity rate is shown overall and stratified by W4SS status. Data are % (n/N).

| | No. expectorated sputa | | | |
|---|---|---|---|---|
| | None[†] (i.e., induction attempted for all sputa) | ≥1[‡] | ≥2[**] | ≥3 (i.e., no induction required) |
| Overall | 31 (49/158) | 69 (109/158) | 64 (101/158) | 62 (98/158) |
| Culture-positive[*] | 20 (10/49) | 16 (17/105) | 15 (15/97) | 14 (14/94) |
| W4SS-negative | 20 (11/55) | 80 (44/55) | 73 (40/55) | 71 (39/55) |
| Culture-positive | 0 (0/11) | 7 (3/44) | 5 (2/40) | 5 (2/39) |
| W4SS-positive | 37 (38/103) | 63 (65/103) | 59 (61/103) | 57 (59/103) |
| Culture-positive | 26 (10/38) | 23 (14/61) | 23 (13/57) | 22 (12/55) |

[*]Four patients with only contaminated cultures excluded from culture-positivity rate calculation.
Number of induced sputum produced: [†]7/49 (14%), 5/49 (10%) and 37/4 (76%) for n=1, n=2 or n=3, respectively; [‡]1/109 (1%), 6/109 (6%) and 0/109 (0%);[**]1/101 (1%), 0/101 (0%) and 0/101 (0%).
Abbreviations: TB, tuberculosis; W4SS, WHO-recommended four-symptom screen.

**Supplementary Table 15**. Yield (proportion of people with at least one positive confirmatory test result detected by a test) and sensitivity and specificity of individual tests according to whether people could expectorate. For each test, yield, sensitivity, and specificity did not differ based on whether a person was able to expectorate sputum, however, this conclusion is limited by the relatively small number of people with a positive test result in the subset of patients with information about their sputum expectoration status. Data are %, 95% CI, and n/N.

| | **All participants** | | **Expectorator** | | **Required induction** | |
|---|---|---|---|---|---|---|
| **Yield** | | | | | | |
| *Sputum* | | | | | | |
| Xpert | 59 (39, 76) 17/29 | | 55 (32, 77) 11/20 | | 67 (30, 93) 6/9 | |
| Ultra | 66 (46, 82) 19/29 | | 65 (41, 85) 13/20 | | 67 (30, 93) 6/9 | |
| Culture | 83 (64, 94) 24/29 | | 80 (56, 94) 16/20 | | 89 (52, 100) 8/9 | |
| *Urine* | | | | | | |
| Ultra* | 34 (18, 54) 10/29 | | 25 (9, 49) 5/20 | | 56 (21, 86) 5/9 | |
| LF-LAM | 24 (10, 44) 7/29 | | 15 (3, 38) 3/20 | | 44 (14, 79) 4/9 | |
| **Sensitivity and specificity** | | | | | | |
| | Sensitivity | Specificity | Sensitivity | Specificity | Sensitivity | Specificity |
| *Sputum* | | | | | | |
| Xpert | 67 (45, 84) 16/24 | 99 (94, 100) 96/97 | 63 (35, 85) 10/16 | 98 (92, 100) 63/64 | 75 (35, 97) 6/8 | 100 (89, 100) 33/33 |
| Ultra | 75 (53, 90) 18/24 | 99 (94, 100) 97/98 | 75 (48, 93) 12/16 | 98 (92, 100) 63/64 | 75 (35, 97) 6/8 | 100 (90, 100) 34/34 |
| *Urine* | | | | | | |
| Conc. Ultra | 32 (14, 55) 7/22 | 100 (96, 100) 84/84 | 21 (5, 51) 3/14 | 100 (94, 100) 57/57 | 50 (16,84) 4/8 | 100 (87, 100) 27/27 |
| LF-LAM | 25 (10, 47) 6/24 | 99 (94, 100) 97/98 | 13 (2, 38) 2/16 | 98 (92, 100) 63/64 | 50 (16,84) 4/8 | 100 (90, 100) 34/34 |

*Conc. and unconc. positive results included.
Abbreviations: CI, confidence interval; Conc., concentrated; LF-LAM, Determine TB LAM Ag, NPV, negative predictive value; PPV, positive predictive value; TB, tuberculosis, Ultra, Xpert MTB/RIF Ultra; Unconc., unconcentrated; W4SS, WHO-recommended four-symptom screen.
